# Predictors of suicidal thoughts and behavior in children: results from penalized logistic regression analyses in the ABCD study

**DOI:** 10.1101/2021.02.15.21251736

**Authors:** Laura S. van Velzen, Yara J. Toenders, Aina Avila-Parcet, Richard Dinga, Jill A. Rabinowitz, Adrián I. Campos, Neda Jahanshad, Miguel E. Rentería, Lianne Schmaal

## Abstract

Despite numerous efforts to predict suicide risk in children, the ability to reliably identify youth that will engage in suicide thoughts or behaviors (STB) has remained remarkably unsuccessful. To further knowledge in this area, we apply a novel machine learning approach and examine whether children with STB could be differentiated from children without STB based on a combination of sociodemographic, physical health, social environmental, clinical psychiatric, cognitive, biological and genetic characteristics. The study sample included 5,885 unrelated children (50% female, 67% white) between 9 and 11 years old from the Adolescent Brain Cognitive Development (ABCD) study. Both parents and youth reported on children’s STB and based on these reports, we divided children into three subgroups: 1. children with current or past STB, 2. children with psychiatric disorder but no STB (clinical controls) and 3. healthy control children. We performed binomial penalized logistic regression analysis to distinguish between groups. The analyses were performed separately for child-reported STB and parent-reported STB. Results showed that we were able to distinguish the STB group from healthy controls and clinical controls (area under the receiver operating characteristics curve (AUROC) range: 0.79-0.81 and 0.70-0.78 respectively). However, we could not distinguish children with suicidal ideation from those who attempted suicide (AUROC range 0.49-0.59). Factors that differentiated the STB group from the clinical control group included family conflict, prodromal psychosis symptoms, impulsivity, depression severity and a history of mental health treatment. Future research is needed to determine if these variables prospectively predict subsequent suicidal behavior.

## Introduction

Despite national and international prevention efforts aimed at reducing suicide risk, the rate of suicide still continues to rise globally [1]. Suicidal thoughts and behaviors typically emerge during adolescence, and their incidence rates rise sharply from childhood to adolescence [2]. Suicide is the second leading cause of death for young people between 10 and 24 years of age [3, 4]. To better target prevention and intervention efforts, we must increase our understanding of risk factors for suicidal thoughts and behaviors in children and adolescents.

To our knowledge, only two studies have investigated risk factors associated with suicidal thoughts and behaviors in a very large sample of children between the ages of 9 and 11 (N=11,875) in the Adolescent Brain Cognitive Development Study [5–7]. DeVille et al. [6] examined social-environmental factors using generalized linear mixed-effects models and revealed that higher levels of family conflict was associated with suicidal ideation, while low parental monitoring was associated with both ideation and attempt. Janiri et al. [5] examined a broader range of potential risk and protective factors for suicidality using logistic regression and also showed that higher levels of family conflict was a risk factor for suicidality, the presence of child psychopathology and longer weekend screen time were also found to be risk factors, while greater parental supervision and positive school involvement were protective factors. However, these studies have not included biological measures (e.g. genetics or regional brain activity), which have been shown to play a role in suicidal thoughts and behavior in adolescents [8]. In addition, these studies have not examined whether a combination of factors, instead of examining associations per risk factor, distinguishes children with and without suicidal thoughts or behavior.

To address these gaps, we examine whether a combination of a broad range of factors (sociodemographic, physical health, clinical psychiatric, cognitive, social environmental, genetic, and biological factors) in a sample of almost 6,000 unrelated children in the ABCD study could differentiate children with a lifetime history of suicidal thoughts and/or suicide attempt and two control groups. Since a large number of children in the suicidal thoughts and behavior (STB) group also have a psychiatric disorder, the control groups were: 1) children without psychiatric disorder (healthy controls; HC), 2) children with psychiatric disorder but no history of suicidal thoughts or behavior (clinical controls; CC). To this end, we used binomial penalized logistic regression and a feature selection approach, which can determine which type of predictors contribute most to the classification of STB.

In addition to examining risk for suicidal thoughts and behaviors, it is important to identify clinical, biological, and cognitive factors that distinguish between individuals who only think about suicide (suicidal ideation), and those who attempt suicide [e.g. 9, 10]. This is relevant as it has been shown that only one third of individuals with suicidal thoughts actually attempt suicide [11]. Thus, identifying factors that differentiate these individuals may identify young people at high risk for suicidal behavior and may further inform targeted prevention and intervention efforts. As a final aim, we examined which factors differentiated children with (a history of) suicidal ideation, but no history of suicidal behavior, and those that have attempted suicide during their lives.

## Methods

### Participants

All data included in this study were collected as part of the Adolescent Brain Cognitive Development (ABCD) study (Annual Release 2.1;https://nda.nih.gov/abcd). Data were drawn from the baseline measurement of the ABCD study, which included data from 11,875 children between the ages of 9 and 11 assessed at 22 sites across the United States. The recruitment method and inclusion and exclusion criteria of the ABCD study are described elsewhere [7]. All adolescents provided assent and their parents provided consent. The Institutional Review Board of the University of California at San Diego approved the study protocol and data collection and is responsible for ethical oversight.

In the current study, we only included unrelated children, leading to a sample size of 9,985 children (see Supplemental Note 1 and Supplemental Figure S1). In addition, 9 children were excluded due to missing sociodemographic data, and 4000 children were excluded due to missing neuroimaging data or excluded due to low quality of neuroimaging data (as suggested by the ABCD team). This resulted in a total sample size of 5885 children for the current analysis.

### Definition of outcome groups

Suicidal thoughts and behaviors (interrupted, aborted or actual suicide attempt) and psychiatric diagnoses were assessed using the child-and parent-reported of the computerized Kiddie Schedule for Affective Disorders and Schizophrenia for DSM-5 (KSADS-5) [12]. As previous findings showed low correspondence between parent- and child-reported STBs in the ABCD sample [5], we created two STB outcome variables, one for each reported (see Supplemental Note 2 for more details on group definitions). Children with comorbid psychiatric disorders were not excluded from the STB group. The definitions for the HC and CC group were the same across parent and child outcome variables, however, due to differences in the STB outcome, the sample of the HC and CC groups also differed.

For the parent-reported STB outcome variable, we created three groups based on the K-SADS-5 diagnostic information: 1) healthy control group (no parent-reported or child-reported psychiatric diagnosis was present and no parent-reported lifetime history of suicidal thoughts and behavior; N=2,415;) 2) clinical control group (a parent-reported or child-reported psychiatric diagnosis was present, but there was no lifetime parent-reported history of suicidal thoughts or behavior; N=2,976); 3) STB group (lifetime parent-reported suicidal thoughts or behavior was reported; N=494).

The child-reported STB outcome variable included the following three groups: 1) healthy control group (no parent-reported or child-reported psychiatric diagnosis was present and no child-reported lifetime history of suicidal thoughts or behavior; N=2,367); 2) clinical control group (a parent-reported or child-reported psychiatric diagnosis was present, but there was no lifetime child-reported history of suicidal thoughts or behavior; N=2,985); 3) STB group (lifetime child-reported suicidal thoughts or behavior was reported; N=528).

In addition, ancillary analyses were performed on the individuals that were in the same group according to both the parent and child outcome variables (see Supplemental Note 3).

For secondary analyses, we created two additional outcome variables to distinguish children with lifetime suicidal thoughts (ideation) from children with a history of suicidal behavior (attempt). The child-reported outcome variable included 461 children with self-reported suicidal ideation but no history of attempt and 67 children with a self-reported history of suicide attempt. The parent-reported suicide ideation and suicide attempt outcome variables included 464 children with suicidal ideation but no history of attempt, and 30 children with a history of suicide attempt (see Supplemental Note 4 for more details on group definitions).

### Risk factors

Seven sociodemographic, 13 physical health, 11 social environmental, 56 clinical psychiatric, 14 cognitive functioning, 88 neuroimaging and five genetic variables were included (Table 1), based on available literature, as predictors of group status (for a detailed overview of all included measures please see Supplemental Note 5 and Supplemental Table S1).

**Table 1.**
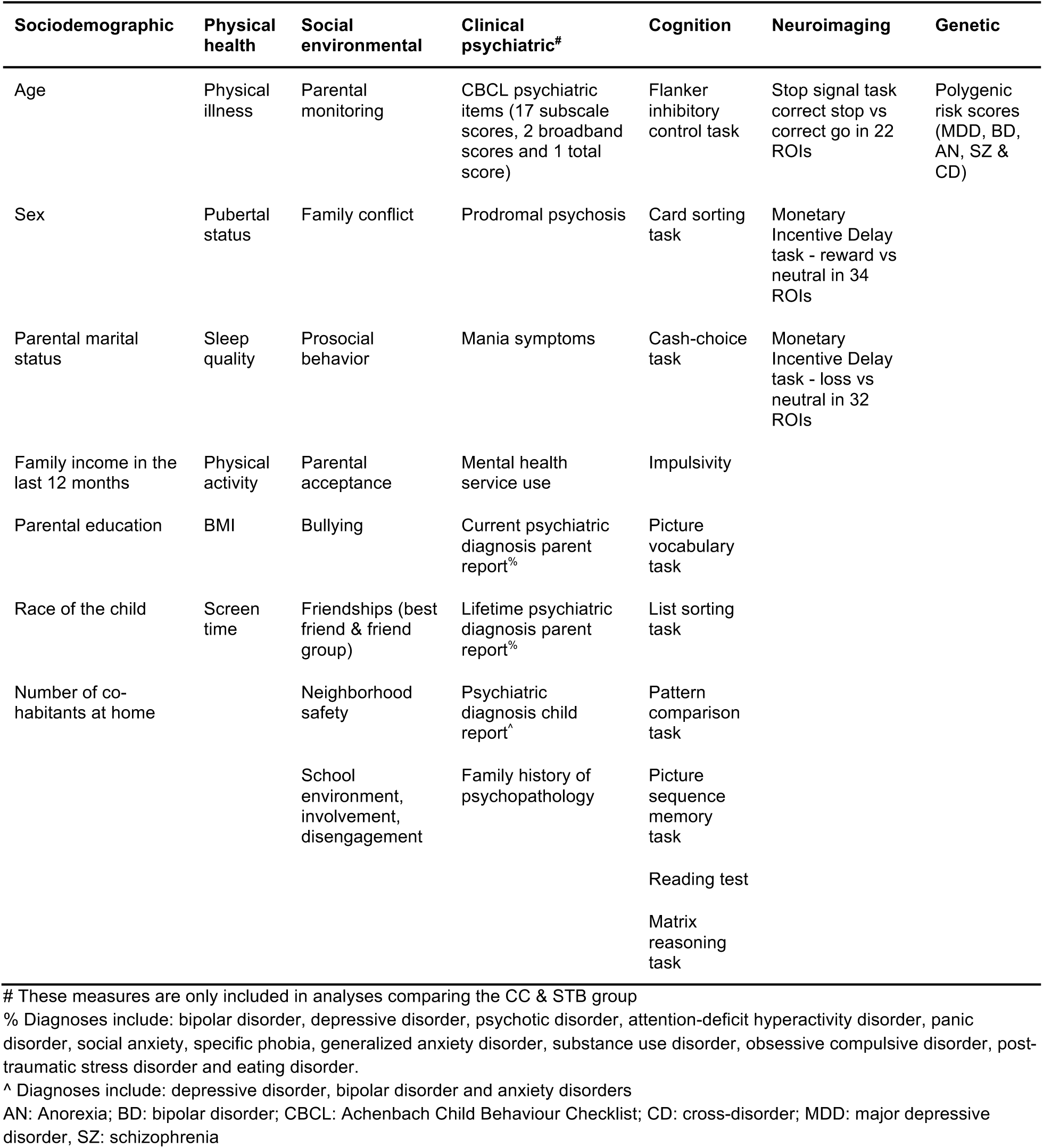
ABCD measures included in this study per domain.

### Statistical analysis

#### Training and independent replication datasets

In order to perform the binomial penalized logistic regression analysis, a training dataset, consisting of 2/3 of the data, and a validation dataset, consisting of 1/3 of the data, were created by randomly splitting the data according to the data collection site to ensure the generalization of model performance to independent sites (see Supplemental Table S2 and Figure 1).

**Figure 1.**
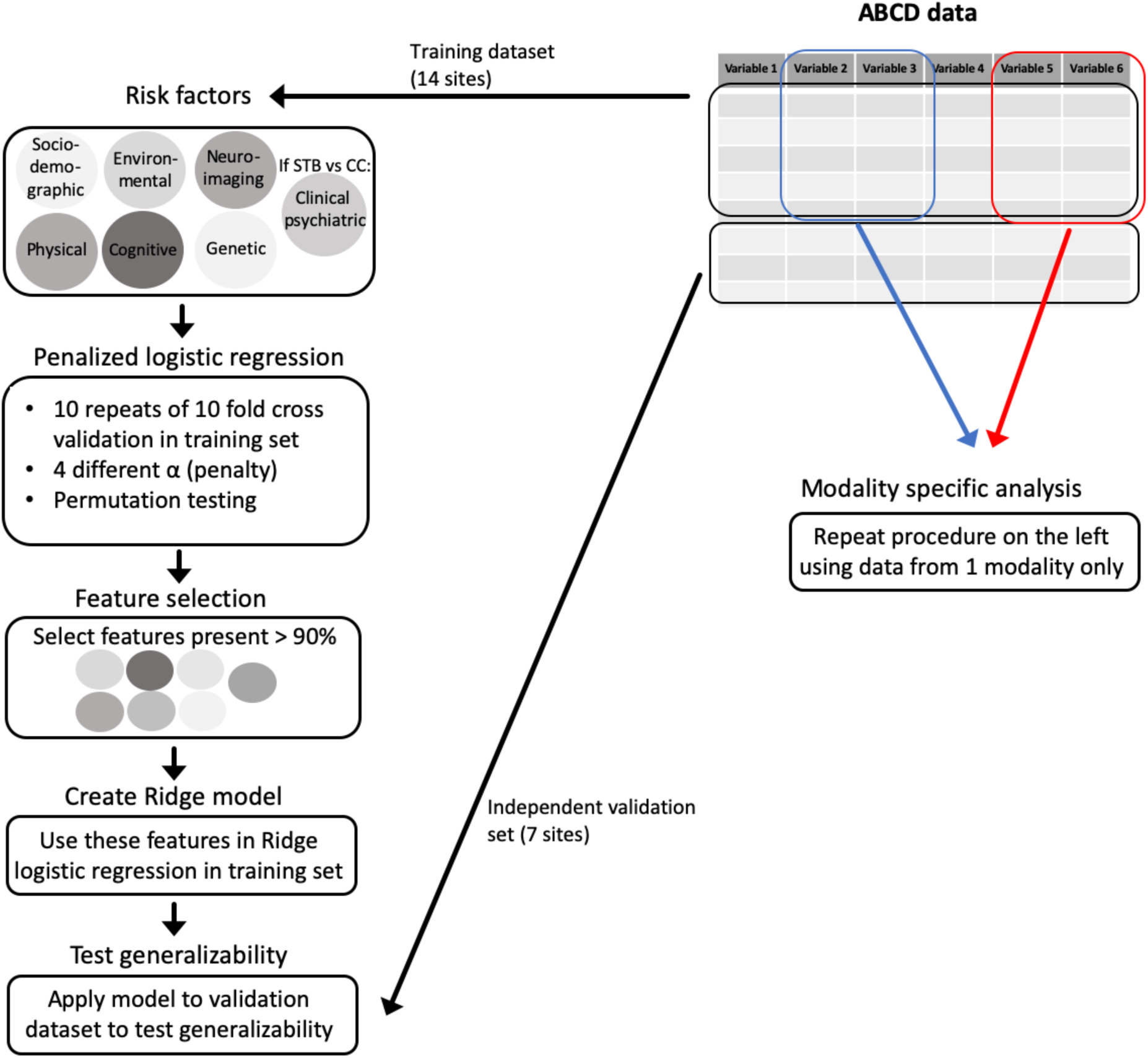
Flowchart to describe the analysis procedure. The ABCD data was split into a training and test set. The training set was used to do a penalized logistic regression in ten-fold cross validation and repeat this ten times with 4 different combinations of the Lasso and Ridge penalty. Features that had a coefficient higher than 0 in 90% or more of the repeats were selected to create a Ridge logistic regression to differentiate groups. This Ridge model was then tested on the test dataset. In addition, the same procedure was repeated only including risk factors from one modality.

#### Classification of group status in the training set

Binomial penalized logistic regression analysis was performed using the package *glmnet* in R [13]. This was applied to a combination of all predictors in the training set to distinguish between (1) the healthy control group; (2) the clinical control group and (3) the STB group. The binomial penalized logistic regression builds a sparse model by adding a penalty which prevents overfitting. This approach combines two types of penalties or regularizations. A Ridge penalty shrinks coefficients, making their contribution to the model small, and a Lasso penalty forces some coefficients to zero, meaning that the feature is not selected for the model. A combination of the two penalties allows for feature selection as well as for features to have a small contribution to the model. Binomial penalized regression was performed with different penalties (alpha levels: 0.25, 0.5, 0.75 and 1), varying between a Lasso penalty (alpha=1) and a combination of Lasso and Ridge penalties (elastic net; alpha’s between 0.25 and 0.75). Ten-fold cross-validation (CV) was applied by dividing the training dataset into 10 sets, and within each CV fold, 9 out of 10 sets were combined to form the training set and 1 was used as the test set. This was repeated 10 times. The glmnet package determined the optimal lambda value by identifying the lambda associated with the minimum Brier score. In each CV fold, we imputed missing values using the caret package [14] in the test set and training set separately, in order to prevent data leakage. Binomial analyses comparing two groups were run (HC vs. CC, CC vs. STB and HC vs STB groups). Binomial analyses were performed instead of multinomial analyses, as a set of clinical psychiatric measures were only non-zero in the CC and STB groups. As the STB group was smaller than the clinical control and healthy control groups, we under-sampled these larger groups within each CV fold to match the size of the STB group.

The performance of the model was assessed using the area under the receiver operating characteristics curve (AUROC). AUROC represents the proportion of times an individual from a positive class (e.g. STB group) is ranked below an individual from a negative class (e.g. HC). In addition, sensitivity, specificity, average of the sensitivity and specificity (accuracy) were calculated. Permutation testing (by comparing the AUROC against the AUROC of the same procedure repeated 1000 with permuted group labels) was used to examine if the model performed significantly above chance level classification. To identify the features that contributed most to the predictive model, the features that had a coefficient of more than 0 in at least 90% of the subsamples were selected.

#### Generalization to the independent validation set

The features that were selected in at least 90% of the subsamples at each alpha (0.25, 0.50, 0.75, 1.00) in the training dataset were used to predict group membership in the independent validation set. This validation set consisted of sites from the ABCD study that were kept separate to ensure independence. This analysis was done to test the generalizability of the predictive model to independent sites and participants. The selected features at each alpha were used in a Ridge logistic regression in the whole training set; this model was then tested on the independent validation set.

#### Modality-specific classification

In order to examine the individual contribution of the different modalities to the classification of the STB groups, we repeated the aforementioned analysis, but only including specific types of predictors, thus performing separate analyses for sociodemographic, physical health, social environmental, clinical psychiatric, cognitive functioning, neuroimaging and genetic predictors.

#### Factors that differentiate ideators from attempters

To examine which factors differentiate between children with a history of attempt from those with suicidal ideation but no history of suicide attempt, the abovementioned binomial penalized logistic regression analysis was performed again with a different outcome variable. For this analysis, the dataset was again divided into a training set and validation set using the same site split as in the main analysis, and the same approach (including the binomial penalized logistic regression with CV, feature selection and Ridge regression) was used to test generalizability; however, only 5 folds were used because the sample size was smaller.

## Results

### Sample characteristics

Age, sex, lifetime psychiatric diagnosis and self-reported suicidal thoughts or behaviors are presented in Table 2 for the three groups (healthy controls, clinical controls, and STB) based on child-reported suicidal thoughts or behaviors. In Table 3, age, sex, lifetime diagnosis and parent-reported suicidal thoughts and behaviors presented for the STB groups based on the parent-reported of suicidal thoughts and behaviors are reported.

**Table 2.** Sample characteristics for the child-reported STB group.

|  | Healthy controls<br>N = 2367 | Clinical controls<br>N = 2985 | Suicidal<br>ideation/attempt<br>N = 528 |
| --- | --- | --- | --- |
| Age (in months) (SD) | 119 (7.44) | 119 (7.38) | 119 (7.52) |
| Sex (% female) | 52.9 % | 47.5 % | 45.1 % |
| Lifetime BD (parent-reported) | NA | 297 (9.9%) | 44 (8.3%) |
| Lifetime BD (child-reported) | NA | 394 (13.2%) | 113 (21.4%) |
| Lifetime DD (parent-reported) | NA | 289 (9.7%) | 62 (11.7%) |
| Lifetime DD (child-reported) | NA | 192 (6.4%) | 74 (14.0%) |
| Lifetime ADHD (parent-reported) | NA | 956 (32.0%) | 154 (29.2%) |
| Lifetime Psychotic disorder (parent-reported) | NA | 120 (4.0%) | 19 (3.6%) |
| Lifetime Panic disorder (parent-reported) | NA | 41 (1.4%) | 10 (1.9%) |
| Lifetime Social anxiety (parent-reported) | NA | 278 (9.3%) | 35 (6.6%) |
| Lifetime Eating disorder (parent-reported) | NA | 549 (18.4%) | 56 (10.6%) |
| Lifetime Specific phobia (parent-reported) | NA | 1413 (47.3%) | 162 (30.7%) |
| Lifetime GAD (parent-reported) | NA | 290 (9.7%) | 60 (11.4%) |
| Lifetime ANX (child-reported) | NA | 127 (4.3%) | 49 (9.3%) |
| Lifetime OCD (parent-reported) | NA | 478 (16.0%) | 87 (16.5%) |
| Lifetime PTSD (parent-reported) | NA | 197 (6.6%) | 60 (11.4%) |
| Lifetime Substance use disorder (parent-reported) | NA | 8 (0.3%) | 1 (0.2%) |
| Lifetime passive suicidal ideation (child-reported) | NA | NA | 386 (73.1%) |
| Lifetime active non-specific suicidal ideation (child-reported) | NA | NA | 275 (52.1%) |
| Lifetime active suicidal ideation + method (child-reported) | NA | NA | 77 (14.6%) |
| Lifetime active suicidal ideation + intent (child-reported) | NA | NA | 35 (6.6%) |
| Lifetime active suicidal ideation + plan (child-reported) | NA | NA | 28 (5.3%) |
| Lifetime preparatory actions toward suicidal behavior (child-reported) | NA | NA | 24 (4.5%) |
| Lifetime interrupted suicide attempt (child-reported) | NA | NA | 8 (1.5%) |
| Lifetime aborted suicide attempt (child-reported) | NA | NA | 28 (5.3%) |
| Lifetime actual suicide attempt (child-reported) | NA | NA | 37 (7.0%) |

Note: ADHD: Attention Deficit Hyperactivity Disorder; ANX: anxiety disorder; BD: bipolar disorder; DD: depressive disorder; GAD: generalized anxiety disorder; OCD: obsessive-compulsive disorder; PTSD: post-traumatic stress disorder; SD: standard deviation

**Table 3.** Sample characteristics for the parent-reported STB group.

|  | Healthy controls<br>N = 2415 | Clinical controls<br>N = 2976 | Suicidal<br>ideation/attempt<br>N = 494 |
| --- | --- | --- | --- |
| Age (in months) (SD) | 119 (7.44) | 119 (7.37) | 120 (7.60) |
| Sex (% female) | 53.0% | 48.4% | 38.5% |
| Lifetime BD (parent-reported) | 0 (0%) | 275 (9.2%) | 69 (14.0%) |
| Lifetime BD (child-reported) | 0 (0%) | 435 (14.6%) | 68 (13.8%) |
| Lifetime DD (parent-reported) | 0 (0%) | 238 (8.0%) | 115 (23.3%) |
| Lifetime DD (child-reported) | 0 (0%) | 218 (7.3%) | 43 (8.7%) |
| Lifetime ADHD (parent-reported) | 0 (0%) | 911 (30.6%) | 205 (41.5%) |
| Lifetime Psychotic disorder (parent-reported) | 0 (0%) | 101 (3.4%) | 38 (7.7%) |
| Lifetime Panic disorder (parent-reported) | 0 (0%) | 33 (1.1%) | 18 (3.6%) |
| Lifetime Social anxiety (parent-reported) | 0 (0%) | 246 (8.3%) | 67 (13.6%) |
| Lifetime Eating disorder (parent-reported) | 0 (0%) | 558 (18.8%) | 52 (10.5%) |
| Lifetime Specific phobia (parent-reported) | 0 (0%) | 1380 (46.4%) | 199 (40.3%) |
| Lifetime GAD (parent-reported) | 0 (0%) | 244 (8.2%) | 107 (21.7%) |
| Lifetime ANX (child-reported) | 0 (0%) | 150 (5.0%) | 26 (5.3%) |
| Lifetime OCD (parent-reported) | 0 (0%) | 461 (15.5%) | 108 (21.9%) |
| Lifetime PTSD (parent-reported) | 0 (0%) | 172 (5.8%) | 86 (17.4%) |
| Lifetime Substance use disorder (parent-reported) | 0 (0%) | 6 (0.2%) | 3 (0.6%) |
| Lifetime passive suicidal ideation (parent-reported) | 0 (0%) | 0 (0%) | 423 (85.6%) |
| Lifetime active non-specific suicidal ideation (parent-reported) | 0 (0%) | 0 (0%) | 232 (47.0%) |
| Lifetime active suicidal ideation + method (parent-reported) | 0 (0%) | 0 (0%) | 59 (11.9%) |
| Lifetime active suicidal ideation + intent (parent-reported) | 0 (0%) | 0 (0%) | 27 (5.5%) |
| Lifetime active suicidal ideation + plan (parent-reported) | 0 (0%) | 0 (0%) | 10 (2.0%) |
| Lifetime preparatory actions toward suicidal behavior (parent-reported) | 0 (0%) | 0 (0%) | 33 (6.7%) |
| Lifetime interrupted suicide attempt (parent-reported) | 0 (0%) | 0 (0%) | 4 (0.8%) |
| Lifetime aborted suicide attempt (parent-reported) | 0 (0%) | 0 (0%) | 6 (1.2%) |
| Lifetime actual suicide attempt (parent-reported) | 0 (0%) | 0 (0%) | 23 (4.7%) |

Note. ADHD: Attention Deficit Hyperactivity Disorder; ANX: anxiety disorder; BD: bipolar disorder; DD: depressive disorder; GAD: generalized anxiety disorder; OCD: obsessive-compulsive disorder; PTSD: post-traumatic stress disorder; SD: standard deviation

### Classification of STB group

#### Classification of STB group: Cross validation model performance

Results of the analysis using the child-reported STB group measures are presented in Table 4. AUROC values were highest when differentiating the HC and STB groups (range: 0.79-0.80 across the different alpha levels), and were lowest for the comparison between HC and CC groups (AUROC range: 0.68-0.69, Figure 2).

**Table 4.**
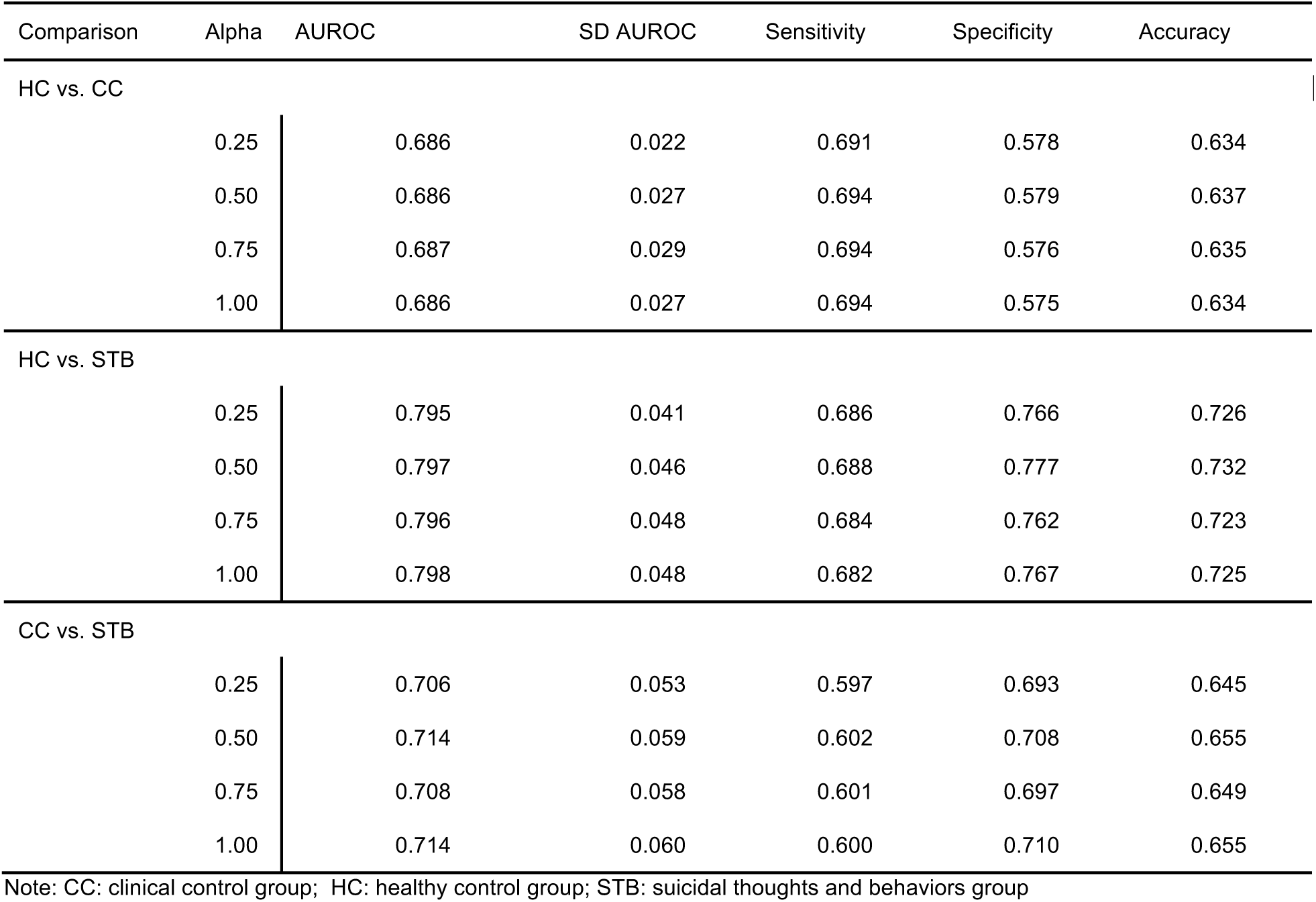
Classification of STB groups (child-reported): Results of binomial penalized logistic regression analysis.

**Figure 2.**
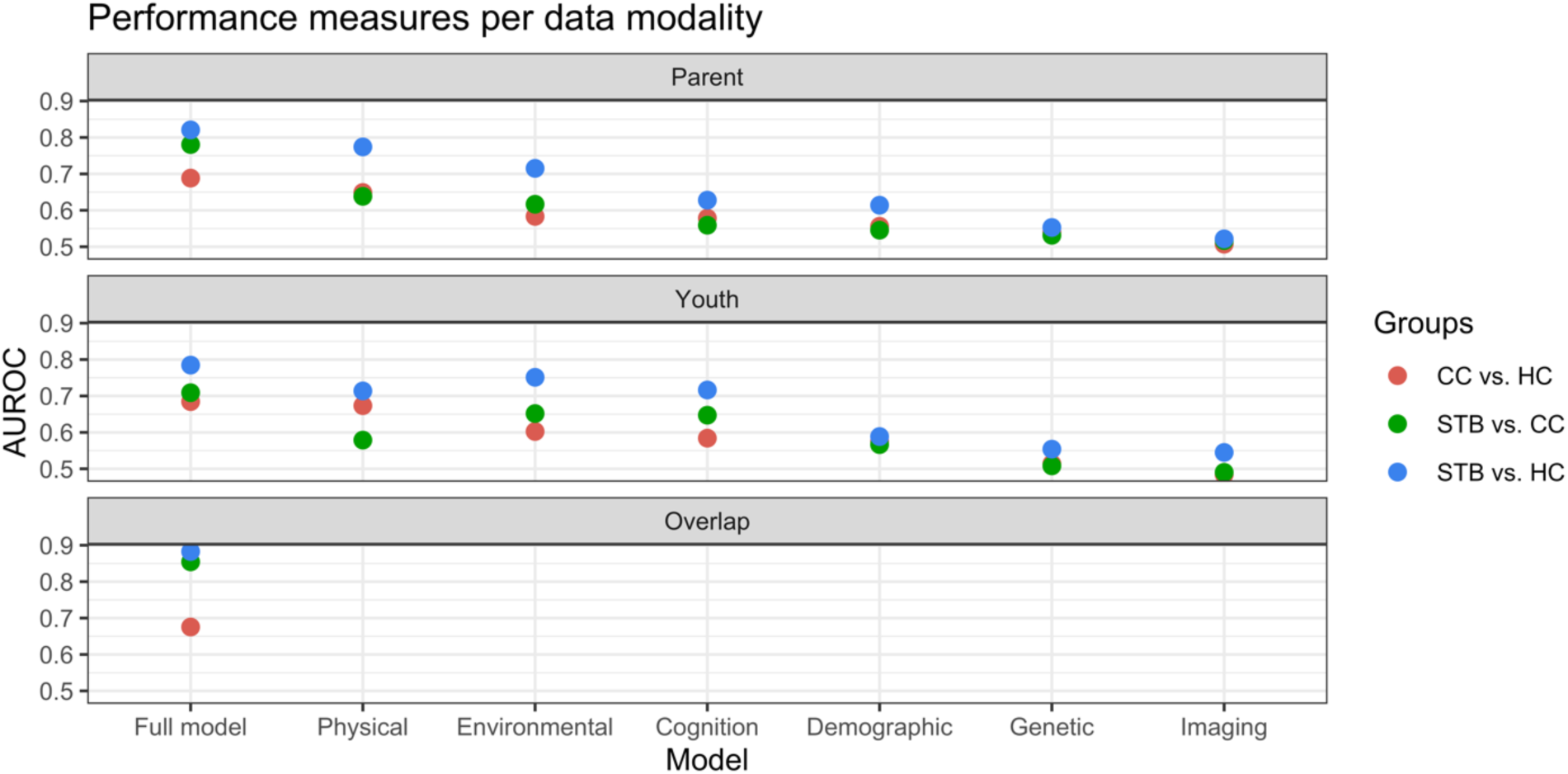
Average AUROC per data modality.

A similar pattern was observed for the results of the analyses using the parent-reported STB group measures (see Table 5), with the highest AUROC observed for the HC vs. STB comparison (range: 0.80-0.81) and lowest for the HC vs. CC comparison (range: 0.68-0.69).

**Table 5.**
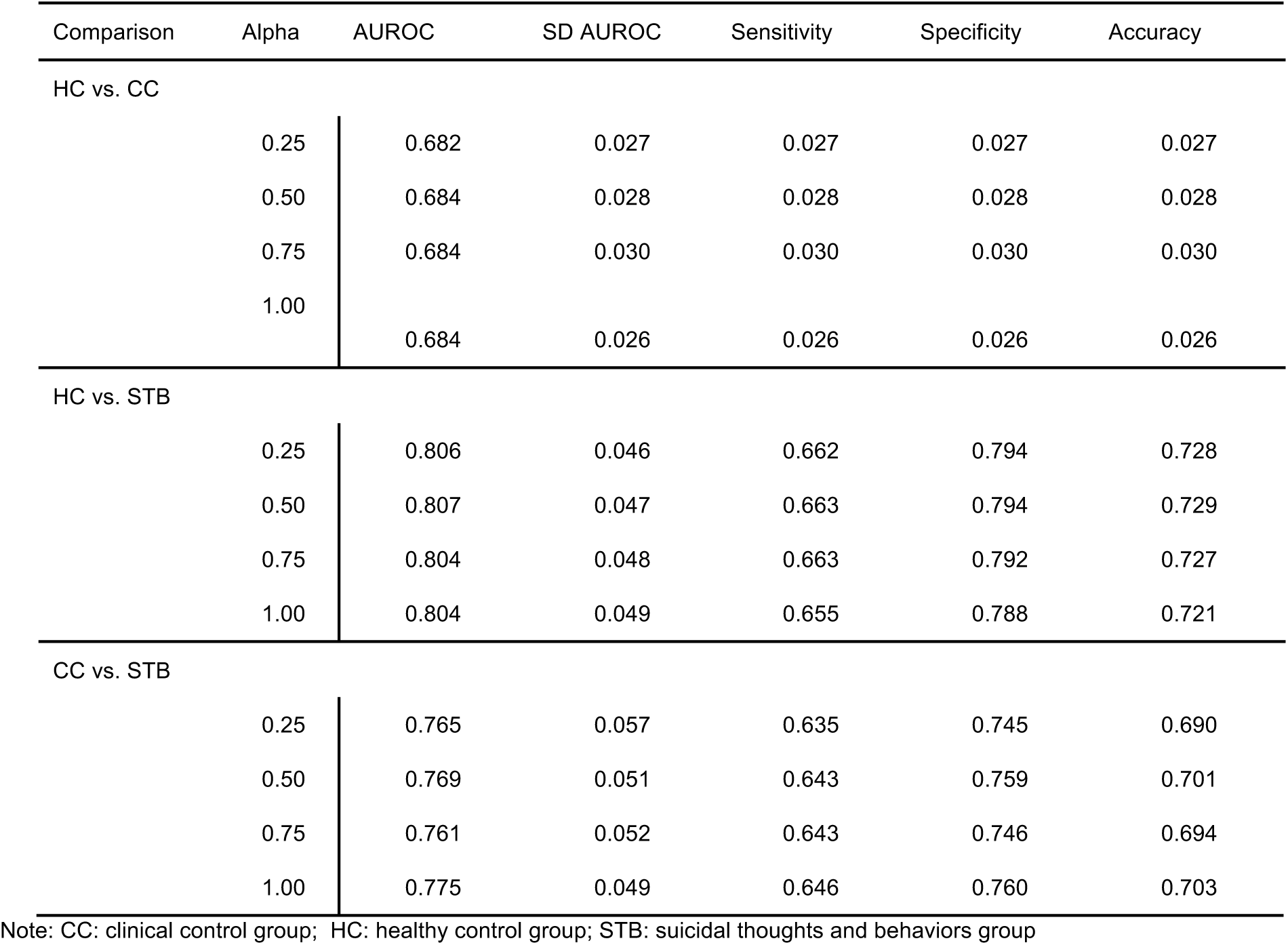
Classification of STB groups (parent-reported): Results of binomial penalized binomial logistic regression analysis.

| Comparison | Alpha | AUROC | SD AUROC | Sensitivity | Specificity | Accuracy |
| --- | --- | --- | --- | --- | --- | --- |
| HC vs. CC |  |  |  |  |  |  |
|  | 0.25 | 0.682 | 0.027 | 0.027 | 0.027 | 0.027 |
|  | 0.50 | 0.684 | 0.028 | 0.028 | 0.028 | 0.028 |
|  | 0.75 | 0.684 | 0.030 | 0.030 | 0.030 | 0.030 |
|  | 1.00 | 0.684 | 0.026 | 0.026 | 0.026 | 0.026 |
| HC vs. STB |  |  |  |  |  |  |
|  | 0.25 | 0.806 | 0.046 | 0.662 | 0.794 | 0.728 |
|  | 0.50 | 0.807 | 0.047 | 0.663 | 0.794 | 0.729 |
|  | 0.75 | 0.804 | 0.048 | 0.663 | 0.792 | 0.727 |
|  | 1.00 | 0.804 | 0.049 | 0.655 | 0.788 | 0.721 |
| CC vs. STB |  |  |  |  |  |  |
|  | 0.25 | 0.765 | 0.057 | 0.635 | 0.745 | 0.690 |
|  | 0.50 | 0.769 | 0.051 | 0.643 | 0.759 | 0.701 |
|  | 0.75 | 0.761 | 0.052 | 0.643 | 0.746 | 0.694 |
|  | 1.00 | 0.775 | 0.049 | 0.646 | 0.760 | 0.703 |

Note: CC: clinical control group; HC: healthy control group; STB: suicidal thoughts and behaviors group

#### Feature selection

Results of the feature selection analysis are presented in Table S4 and S5 for the child-reported STB groups and parent-reported STB groups, respectively. While the same predictors were included in both analyses, the factors that distinguished the child-reported STB group from the clinical controls were race, family conflict, prodromal psychotic symptoms, impulsivity (UPPS-P negative urgency and lack of planning subscales) and the CBCL depression subscale score. The factors that differentiated the clinical controls from the parent-reported STB group included the CBCL depression subscales (anxious depression, DSM5 depression), CBCL conduct disorder subscale score, CBCL internalizing and externalizing broad band scores and a history of mental health treatment.

#### Generalization to the independent validation dataset

The AUROC in the independent validation dataset (separate ABCD sites) using the most contributing features selected (see above), was in line with the AUROCs achieved in the training dataset (see Supplemental Tables S6 and S7). Classifying HC vs. STB, the AUROC ranged between 0.78-0.80 using the child-reported STB group measure and between 0.80-0.82 using the parent-reported STB group measure, using the features that were selected in the training dataset at different alphas. Classifying HC vs. CC, the AUROC ranged between 0.70-0.71 using the child-reported group measure and between 0.70-0.71 using the parent-reported measure. Finally, classifying CC vs. STB, the AUROC ranged between 0.68-0.72 when using the child-reported measure and between 0.70-0.71 when using the parent-reported measure.

#### Modality-specific classification

Results of these analyses are presented in Table S8 and S9 for the child-reported and parent-reported STB group analyses, respectively. For both the classification of the child- and parent reported STB group status, the clinical psychiatric (AUROC range child-reported: 0.67-0.69; parent-reported: 0.76-0.79), physical health (AUROC range child-reported: 0.58-0.73; parent-reported: 0.63-0.78), cognitive functioning (AUROC range child-reported: 0.58-0.72; parent-reported: 0.54-0.65) and social environmental factors (AUROC range child-reported: 0.60-0.74; parent-reported: 0.60-0.71) best predicted STB group status, in contrast to neuroimaging (AUROC range child-reported: 0.49-0.54; parent-reported: 0.50-0.53), sociodemographic measures (AUROC range child-reported: 0.53-0.59; parent-reported: 0.53-0.62) and genetic characteristics (AUROC range child-reported: 0.51-0.57; parent-reported: 0.52-0.56). Similar to the aforementioned results, the highest AUROC values were observed for the HC vs. STB comparison.

### Classification of ideators versus attempters

Results of the analysis used to classify child-reported suicidal ideation versus suicidal attempt are presented in Table 6. AUROC values varied between 0.54 and 0.59 across the different alpha levels. Results of the same analysis, but using the parent-reported group measure showed similar results (AUROC range: 0.49-0.54; see Table 7). As the results show that it is not possible to distinguish these two groups, no further feature selection or modality specific classification was performed.

**Table 6.** Classification of ideation versus suicidal behavior (child-reported): Results of binomial penalized logistic regression analysis.

| Comparison | Alpha | AUROC | SD AUROC | Sensitivity | Specificity | Accuracy |
| --- | --- | --- | --- | --- | --- | --- |
| Ideation versus suicidal behavior |  |  |  |  |  |  |
|  | 0.25 | 0.589 | 0.148 | 0.655 | 0.468 | 0.562 |
|  | 0.50 | 0.566 | 0.127 | 0.685 | 0.408 | 0.546 |
|  | 0.75 | 0.547 | 0.120 | 0.712 | 0.368 | 0.540 |
|  | 1.00 | 0.567 | 0.126 | 0.718 | 0.373 | 0.546 |

**Table 7.** Classification of ideation versus suicidal behavior (parent-reported): Results of binomial penalized logistic regression analysis.

| Comparison | Alpha | AUROC | SD AUROC | Sensitivity | Specificity | Accuracy |
| --- | --- | --- | --- | --- | --- | --- |
| Ideation versus suicidal behavior |  |  |  |  |  |  |
|  | 0.25 | 0.493 | 0.201 | 0.645 | 0.353 | 0.499 |
|  | 0.50 | 0.520 | 0.199 | 0.724 | 0.305 | 0.514 |
|  | 0.75 | 0.513 | 0.156 | 0.674 | 0.318 | 0.496 |
|  | 1.00 | 0.532 | 0.231 | 0.679 | 0.329 | 0.504 |

## Discussion

In a large sample of almost 6,000 unrelated children, we examined whether a combination of different predictors (sociodemographic, physical health, clinical psychiatric, cognitive, psychosocial, neuroimaging and genetic factors) could differentiate healthy children, children with psychiatric disorder but no history of suicidal thoughts or behavior, and children with a lifetime history of suicidal thoughts or suicide attempt (the STB group). Binomial penalized logistic regression analysis showed that the STB group could be distinguished from the HC and CC groups (AUROC range 0.79-0.81 and 0.70-0.78 respectively), but the ability to differentiate the CC and HC group was less accurate (AUROC range 0.68-0.69). These results may be explained by the fact that the children with most severe psychiatric symptoms may have been included in the STB group, thereby reducing the differences between the CC and HC groups. Our model generalized to independent data (separate ABCD recruitment sites (AUROC range 0.68-0.82)). The analyses for groups based on parent- and child-reported measures were performed separately, as a recent study [5] showed low correspondence between parent-reported and child-reported measures of suicidal thoughts and behaviors in the ABCD study. The AUROCs of these analyses were very similar, as we were able to distinguish the groups based on both the parent- and child-reported measures. Children with a lifetime history of suicidal ideation could not be distinguished from those with a lifetime history of suicide attempt (AUROC range 0.49-0.59).

The classification of STB and identifying contributing risk factors are important aims in suicide research as it may help identify those at risk and help target prevention and intervention efforts. We were able to classify the STB group from the CC group with sensitivity between 0.59 and 0.65, and specificity between 0.69 and 0.77. This means that 2 out of 5 children with suicidal thoughts or behavior would be missed, and 1-2 out of 5 children would be considered to have suicidal thoughts or behavior when they do not. This sensitivity is lower than the sensitivity of existing suicide scales in predicting suicide attempt [15, 16], and not yet sufficient to be used as a clinical decision tool. Our findings are in line with three meta-analyses that showed that (a combination of) psychological or biological measures were limited in their ability to predict suicide or suicidal behavior [16–18] showing that classification of suicidal thoughts and behaviors is complex, and adding to the current debate around precision medicine in suicide research [e.g. 19].

When the risk factors were divided into separate modalities to examine their unimodal predictive characteristics, the AUROC values for social environmental, physical, cognitive, and clinical psychiatric modalities were higher than the AUROC values for neuroimaging, genetic and sociodemographic modalities. This finding was in line with the strongest contributing features when all predictors were combined in one analysis, as these features were mainly from the social environmental, and clinical psychiatric categories. The fMRI-based measures included did not seem to contribute to the classification of children with STB In contrast to these findings, previous studies have found that functional brain alterations in the prefrontal cortex are related to STB [20, 21] and contribute to the classification of suicidal youth [22]. However, our findings are consistent with a neuroimaging-specific evaluation of this same cohort, in which no association was found between suicidal thoughts and behaviors and functional neuroimaging measures [23]. These discrepant findings between ABCD and other studies could potentially be explained by the younger age of participants in the ABCD study, the fact that ABCD is a population study, or methodological issues that have been described elsewhere [24].

Most features that contributed to the model classifying HC and STB also contributed when classifying CC from HC. When the STB group was differentiated from the CC group, family conflict, prodromal psychosis, severity of mental health symptoms and measures of impulsivity were amongst the features that contributed most to the model’s predictions. These findings highlight the potential need for clinicians to consider alternative interventions, including family-based psychological interventions to decrease family conflict [25] or neuropsychological training to increase cognitive control and planning abilities; and emotional regulation skills, distress tolerance training or mindfulness based interventions in order to decrease negative urgency and modulate impulsivity in suicidal individuals. Surprisingly, parent-reported child mental health service use, predicted parent-reported STB, but not child-reported STB, further highlighting the low correspondence between parent- and child-reported STB.

Understanding which children will experience suicidal thoughts or attempt suicide has important implications for suicide prevention and clinical practice [26]. In this cross-sectional study, we were unable to differentiate children with suicidal thoughts from children with a history of suicidal behavior, potentially suggesting a shared etiology between ideation and attempt in this age group. A large study in 16 year-olds showed that, compared to adolescents with suicidal thoughts, those that attempted suicide were more often exposed to self-harm by friends or family members, were more likely to be diagnosed with a psychiatric disorder, more often were female, exposed to trauma, more impulsive and had specific personality characteristics (i.e. high sensation seeking and low conscientiousness) [27]. A second large study conducted among adolescents and young adults showed that acquired capability, impulsivity, mental imagery about death and exposure to suicidal behavior were more common in those who attempted suicide compared to ideators [10]. Meta-analyses showed that traumatic life events, history of abuse, drug use disorders, and alterations in decision making and impulsivity were more common in attempters than ideators, while depression, alcohol use, hopelessness, and sociodemographic variables did not differ between attempters and ideators [28, 29]. We were unable to include most of the aforementioned variables in our logistic regression model, which may explain why our classification performance was poorer than that observed in previous studies. The variables that contributed to the classification of STB from CC, were unable to distinguish ideation from an attempt, as they may be related to suicidal thoughts and behavior in general, and do not differ between ideators and attempters. In addition, only 67 children reported a history of suicide attempt and only 30 parents reported that their child had a history of suicide attempt, which may have limited our power to detect small effects. Finally, the young age of these participants may have added additional noise to the classification, as a larger fraction of the ideation group may attempt suicide in the future compared to studies with older participants.

This is the first study to combine multimodal features to classify children with suicidal thoughts and behavior from the CC and HC participants in the ABCD study, and builds on previous work by Janiri et al. and DeVille et al. [5, 6]. Strengths of this study include the large sample size of unrelated participants, the availability of many different types of predictors, including clinical, sociodemographic, biological and cognitive measures, and the use of an ecologically valid control group consisting of children with a psychiatric disorder. An additional strength is rigorous validation using cross validation and an independent out of sample validation which avoids overly optimistic results due to overfitting in the training set. The findings need to be interpreted in the light of a few limitations, including the cross-sectional nature of the data. Longitudinal data collection for participants enrolled in the ABCD study is planned at 2 year intervals for a total of 10 years, and future studies may build on these baseline models to predict suicidal thoughts or behavior throughout adolescence. Finally, no measures of the severity or frequency of suicidal ideation or behavior were available, which limited our ability to examine specific subgroups with varying suicidal severity.

Identifying factors that differentiate children with psychiatric disorder with and without a history of suicidal thoughts or behavior can inform risk assessments and identify new treatment targets. Our study shows that social environment (family conflict), cognitive (impulsivity) and clinical measures (e.g. severity of prodromal psychosis symptoms, severity of depression) differentiate children with and without a history of suicidal thoughts and behavior. More studies in a larger sample of attempters are needed to confirm whether the factors identified in our study differentiate those with ideation from those with a history of attempt and prospectively predict subsequent suicidal behavior. In addition, future studies could determine whether including additional variables (e.g., trauma-related variables or suicide-related measures) improves classification. This work highlights the need for clinicians to monitor children who present multiple risk factors and may inform future socio-environmental interventions that may contribute to suicide prevention in at-risk children.

## Data Availability

Data used in the preparation of this article were obtained from the Adolescent Brain Cognitive DevelopmentSM(ABCD) study (https://abcdstudy.org), held in the NIMH Data Archive (NDA).

## Acknowledgements

This work was supported by the MQ Brighter Futures Award MQBFC/2 (LS) and the National Institute of Mental Health of the National Institutes of Health under Award Number R01MH117601 (LS, NJ). LS is supported by a NHMRC Career Development Fellowship (1140764).

Data used in the preparation of this article were obtained from the Adolescent Brain Cognitive Development^SM^(ABCD) study (https://abcdstudy.org), held in the NIMH Data Archive (NDA). This is a multisite, longitudinal study designed to recruit more than 10,000 children age 9-10 and follow them over 10 years into early adulthood. The ABCD Study® is supported by the National Institutes of Health and additional federal partners under award numbers U01DA0401048, U01DA050989, U01DA051016, U01DA041022, U01DA051018, U01DA051037, U01DA050987, U01DA041174, U01DA041106, U01DA041117, U01DA041028, U01DA041134, U01DA050988, U01DA051039, U01DA041156, U01DA041025, U01DA041120, U01DA051038, U01DA041148, U01DA041093, U01DA041089, U24DA041123, U24DA041147. A full list of supporters is available at https://abcdstudy.org/federal-partners.html. A listing of participating sites and a complete listing of the study investigators can be found at https://abcdstudy.org/consortium_members/. ABCD consortium investigators designed and implemented the study and/or provided data but did not necessarily participate in analysis or writing of this report. This manuscript reflects the views of the authors and may not reflect the opinions or views of the NIH or ABCD consortium investigators. The ABCD repository grows and changes over time. The ABCD data used in this report came from 10.15154/1520786. DOIs can be found at nda.nih.gov.

