## Supplemental material for "Predictors of suicidal thoughts and behavior in children: results from penalized logistic regression analyses in the ABCD study"

### Supplemental Materials

### Supplemental Figure S1. Sample selection and group sample sizes

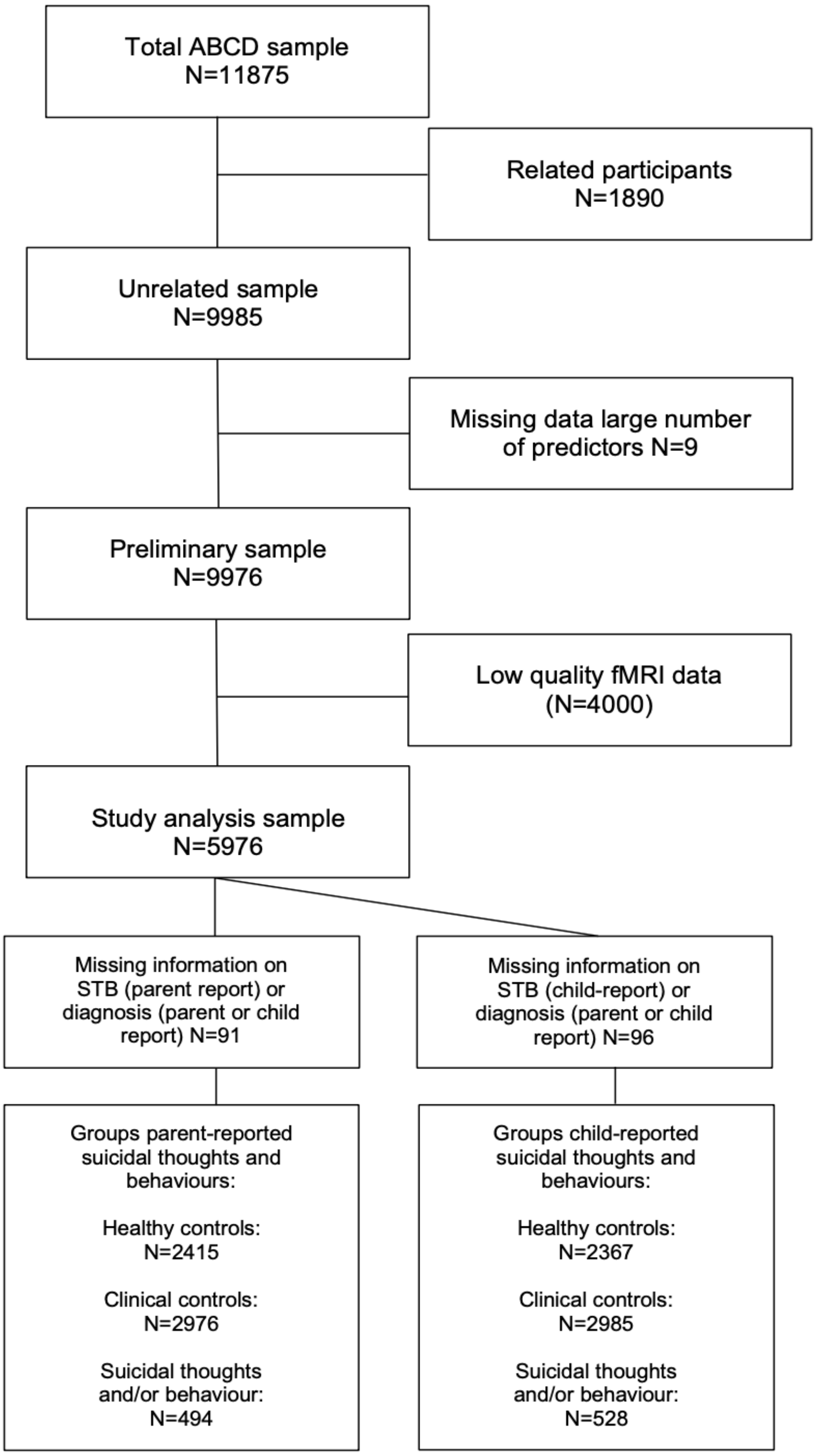

**Supplemental Note 1: Selection of unrelated participants**

The ABCD study includes a total of 11,875 children, some of whom are siblings. In the current study, we chose to include only unrelated participants. Instead of randomly selecting a sibling from a family, we chose to select siblings based on STB using the selection strategy described below to maximize the sample size of the STB groups.

In particular, if one of the siblings had a lifetime history of suicidal behavior (actual, interrupted or aborted attempt according to the parent- or child reported), this sibling was selected from that family. If in a family, multiple siblings had a lifetime history of suicidal behavior, we selected the sibling that had a history of an actual attempt over siblings with an aborted or interrupted attempt.

If none of the siblings had a lifetime history of suicidal behavior, we examined lifetime history of suicidal ideation (parent- or child-reported). If multiple siblings had a history of suicidal ideation, we selected the sibling with the most severe ideation (coded as: 5: active ideation with a plan, 4: active ideation with intent, 3: active ideation with a specific method, 2: active non-specific ideation, 1: passive ideation). If there were multiple siblings with similar severity of suicidal ideation, one sibling was selected randomly.

If none of the siblings had a lifetime history of suicidal behavior or suicidal ideation, one sibling was then selected randomly.

The sample of selected siblings was then combined with the sample of participants who did not have siblings included in the ABCD study, resulting in a total of 9985 participants.

**Supplemental Note 2: STB outcome group definition**

Previous findings showed low correspondence between parent- and child reported STB [5], therefore we created two STB outcome variables: the parent-reported STB group variable and the child-reported STB group variable.

<u>Child-reported STB group variable</u>

The child reported STB group variable consists of three groups:

1. STB group (N=528)
2. Clinical control group (N=2985)
3. Healthy control group (N=2367)

The STB group includes children who reported lifetime suicidal ideation or suicidal behavior, which means that they endorse one or more of the following measures from the K-SADS-5: present or past passive suicidal ideation (ksads_23_946_t, ksads_23_957_t), present or past active non-specific suicidal ideation (ksads_23_947_t, ksads_23_958_t), present or past active suicidal ideation with a specific method (ksads_23_948_t, ksads_23_959_t), present or past active suicidal ideation with intent (ksads_23_949_t, ksads_23_960_t), present or past active suicidal ideation with a plan (ksads_23_950_t, ksads_23_961_t), present or past preparatory actions toward imminent suicidal behavior (ksads_23_951_t, ksads_23_962_t), present or past interrupted suicide attempt (ksads_23_952_t, ksads_23_963_t), present or past aborted suicide attempt (ksads_23_953_t, ksads_23_964_t), present or past actual suicide attempt (ksads_23_954_t, ksads_23_965_t).

The clinical control group did not reported lifetime suicidal ideation or suicidal behavior, but did have a lifetime psychiatric diagnosis based on parent- or child-reported of the K-SADS-5 of one or more of the following disorders: bipolar disorder, depressive disorder, psychotic disorder, ADHD, panic disorder, social anxiety disorder, specific phobia, generalized anxiety disorder, substance use disorder, obsessive-compulsive disorder, alcohol disorder, PTSD, eating disorder. As the available K-SADS-5 diagnoses based on child-reported are very limited, we determined there was a diagnosis if this was derived from the parent-reported or the child-reported.

The healthy control group includes children who have not reported lifetime history of suicidal thoughts or behaviors, and who do not have a lifetime diagnosis of a psychiatric disorder according to the K-SADS-5 parent- and child-reports.

Parent-reported STB group variable

The parent-reported STB group variable consists of three groups:

1. STB group (N=494)
2. Clinical control group (N=2976)
3. Healthy control group (N=2415)

Here, the STB group includes children whose parents reported that their child had a lifetime history of suicidal ideation or suicidal behavior, which includes the following measures from the K-SADS-5 parent-reported: present or past passive suicidal ideation (ksads_23_946_p, ksads_23_957_p), present or past active non-specific suicidal ideation (ksads_23_947_p, ksads_23_958_p), present or past active suicidal ideation with a specific method (ksads_23_948_p, ksads_23_959_p), present or past active suicidal ideation with intent (ksads_23_949_p, ksads_23_960_p), present or past active suicidal ideation with a plan (ksads_23_950_p, ksads_23_961_p), present or past preparatory actions toward imminent suicidal behavior (ksads_23_951_p, ksads_23_962_p), present or past interrupted suicide attempt (ksads_23_952_p, ksads_23_963_p), present or past aborted suicide attempt (ksads_23_953_p, ksads_23_964_p), present or past actual suicide attempt (ksads_23_954_p, ksads_23_965_p).

The clinical control group did not have a history of suicidal thoughts and behavior according to the K-SADS-5 parent report, but had a lifetime diagnosis of psychiatric disorder (parent-reported or child-reported), similar to what is described above for the child-reported STB group).

The healthy control group does not have a history of suicidal thoughts and behavior according to the K-SADS-5 parent reported and does not have a lifetime diagnosis of psychiatric disorder according to the K-SADS-5 parent or child-reported.

**Supplemental Note 3: Overlapping outcome groups**

The child-reported and parent-reported outcome groups differed from each other, therefore the analyses were repeated using only the individuals that were in the same group according to the K-SADS-5 child and parent report (the overlapping sample). The overlapping outcome variable included the following three groups: 1) healthy control (N=2,298), 2) clinical control group (N=2,693), 3) STB group (N=139). The results in the overlapping sample are presented in Table S3.

**Supplemental Note 4: Ideation vs suicidal behavior group definition**

Child-reported Ideation vs. suicidal behavior group variable

The child-reported Ideation vs. attempt group variable consists of two groups:

1. Lifetime self-reported suicidal ideation, but no self-reported history of attempt group (N=461)
2. Self-reported history of suicidal behavior group (N=67)

The suicidal behavior group includes children who endorse one or more of the following KSADS-5 items: present or past interrupted suicide attempt (ksads_23_952_t, ksads_23_963_t), present or past aborted suicide attempt (ksads_23_953_t, ksads_23_964_t), present or past actual suicide attempt (ksads_23_954_t, ksads_23_965_t).

Children in the suicidal ideation group did not report past or present interrupted, aborted or actual suicide attempts, but endorsed one or more of the following items on suicidal ideation: present or past passive suicidal ideation (ksads_23_946_t, ksads_23_957_t), present or past active non-specific suicidal ideation (ksads_23_947_t, ksads_23_958_t), present or past active suicidal ideation with a specific method (ksads_23_948_t, ksads_23_959_t), present or past active suicidal ideation with intent (ksads_23_949_t, ksads_23_960_t), present or past active suicidal ideation with a plan (ksads_23_950_t, ksads_23_961_t).

Parent-reported Ideation vs. suicidal behavior group variable

The parent-reported Ideation vs. attempt group variable consists of two groups:

1. Lifetime parent-reported suicidal ideation, but no parent-reported history of attempt group (N=464)
2. Parent-reported history of suicidal behavior group (N=30)

Here, the suicidal behavior group includes children whose parents reported that their child hasda lifetime history of suicidal behavior, which included the following measures from the K-SADS-5 parent-reported: present or past interrupted suicide attempt (ksads_23_952_p, ksads_23_963_p), present or past aborted suicide attempt (ksads_23_953_p, ksads_23_964_p), present or past actual suicide attempt (ksads_23_954_p, ksads_23_965_p).

The suicidal ideation group did not have a history of aborted, interrupted of actual suicide attempts according to their parents, but their parents endorsed one or more of the following KSADS items on suicidal ideation: present or past passive suicidal ideation (ksads_23_946_p, ksads_23_957_p), present or past active non-specific suicidal ideation (ksads_23_947_p, ksads_23_958_p), present or past active suicidal ideation with a specific method (ksads_23_948_p, ksads_23_959_p), present or past active suicidal ideation with intent (ksads_23_949_p, ksads_23_960_p), present or past active suicidal ideation with a plan (ksads_23_950_p, ksads_23_961_p).

**Supplemental Note 5: Included features per dimension**

*Sociodemographic factors*

Age at time of assessment (in months), birth sex, parent-reported race of the child (White, Black, Native-American, Asian, other or mixed race), marital status of the parents, combined educational level of both parents, total family income (as a measure of socio-economic status) and the number of cohabitants in the household were determined using questions from the PhenX toolkit (parent-reported; [30]) and included as sociodemographic predictor variables.

*Physical factors*

The included physical factors were pubertal development, sleep disturbance, physical activity, BMI, screen time and physical illness. Pubertal status (child-reported) was assessed using the Pubertal Development Scale [31]. Total scores from the Sleep Disturbance Scale for Children [32, 33] (parent-reported) and its subscales (sleep breathing disorders, disorders of sleep arousal, sleep-wake transition disorders, disorders of excessive somnolence and sleep hyperhydrosis) were included as measures of sleep function. Body mass index (BMI) was calculated based on weight and height. Exercise was assessed by questions from the Youth Risk behavior Survey [34], by asking the child how many days in the past week they were active for at least an hour and how many days they did exercises to strengthen their muscles. Physical illness was included as a dichotomous variable, which was scored as 'yes' when parents reported the child had (a history of) brain injury, cancer, cerebral palsy, diabetes, epilepsy, multiple sclerosis or heart disease in the Magic Health Services Utilization Questionnaire [35]. Finally, screen time was assessed by asking children how many hours they typically spend during a weekday and a weekend day on a computer, tablet, cellphone or other electronic device [36].

*Social environment factors*

The social environmental factors included parental monitoring, family conflict, prosocial behavior, parental acceptance, bullying, friendship, neighborhood safety and school environment. Friendship was assessed by asking parents if their child has a best friend and if their child has a group of friends in the introduction to the KSADS-5 [12]. Bullying at school or in the neighborhood was assessed using the parent-reported of the introduction to the KSADS-5 [12]. Neighborhood safety/crime was assessed by calculating the mean of the parent-reported Neighborhood Safety/Crime Survey [37]. While bullying, friendships and neighborhood safety were available from the parent- and child-reported, the parent-reported was used here due to the high number of missing values in the child-reported bullying measure and because more detailed information was available on neighborhood safety and friendships in the parent-reported. Family conflict was assessed with the family conflict subscale score of the Family Environment Scale (child-reported) [38]. Prosocial behavior was measured using the mean of the prosocial behavior subscale of the Strengths and Difficulties Questionnaire (child-reported) [39]. Child-reported parental acceptance was assessed using the acceptance subscale from the short version of the Children's report of Parental behavior Inventory (CRPBI-short) [40]. Parental monitoring was determined with the mean of the child-reported Parental Monitoring Questionnaire [41]. School environment, involvement and disengagement were assessed with the child-reported School Risk and Protective Factors Survey [42].

*Clinical psychiatric factors*

Psychopathology, prodromal psychosis, mania symptoms, mental health service use, current and lifetime psychiatric diagnosis and family history of psychopathology were included as clinical psychiatric factors. As the scores on clinical psychiatric measures were very low and by definition no psychiatric diagnoses were present in the HC group, the clinical psychiatric measures described below (with the exception of the family history of mental health issues) were only included in the binomial penalized logistic regression models when comparing the clinical controls and individuals with suicide thoughts or behavior.

Dimensional psychopathology was assessed using the T-scores of the 20 subscales of the Achenbach Child behavior Checklist (CBCL; parent-reported) [43]. Mental health service use was determined by asking parents if their child ever received treatment for mental health issues or addiction and what type of treatment was received (e.g. outpatient care, inpatient care) in the Introduction to KSADS-5 [12]. Family history of mental health issues was determined using a modified version of the Family History Assessment from NCANDA [44] (parent-reported), and was coded in this study as a dichotomous variable, with 'yes' indicating a family history of mania, psychosis, nerves, mental health hospitalization or mental health treatment. Dimensional mania symptoms of the child were assessed using the ten-item parent-reported Mania Scale from the Parent General behavior Inventory (PGBI) [45]. Child-reported psychotic symptoms were assessed using the brief version of the Pediatric Psychosis Questionnaire [46]. Current and past psychiatric diagnoses were determined from the KSADS-5 parent-reported and child-reported.

*Cognitive factors*

Age-corrected standardized scores (one measure per test) were obtained from the NIH Toolbox picture vocabulary test, flanker inhibitory control and attention test, list sorting working memory test, dimensional change card sorting test, pattern comparison processing speed test, picture sequence memory test and oral reading recognition test (http://www.nihtoolbox.org). In addition, the total score of the WISC-V Matrix Reasoning scale [47] and the Cash Choice Task delay of gratification measure [48] were included. Finally, total scores on the five subscales of the self-reported UPPS-P scale (negative urgency, lack of planning, sensation seeking, positive urgency and perseverance) were included as measures of impulsivity [49, 50].

*Task-based neuroimaging measures*

Participants underwent a functional magnetic resonance imaging (MRI) scan. The neuroimaging protocol, scan acquisition parameters, and pre-processing procedures are described in detail elsewhere [51]. For the current study, we included derived brain activity measures from various regions of interest during the Stop Signal Task (which measures response inhibition) and the Monetary Incentive Delay Task (which measures reward monitoring and processing) as predictors in our model. Details about these tasks can be found in [51].

For the Stop Signal Task, we included regionally averaged beta-estimates from 14 cortical and 8 subcortical regions of interest during the correct stop versus correct go condition (see

Supplemental Table S1). These regions of interest were selected based on findings from previous meta-analyses [52, 53].
For the Monetary Incentive Delay task we included beta weights for reward anticipation from 22 cortical and 12 subcortical regions during the reward versus neutral condition, and beta weights for loss anticipation from 18 cortical and 14 subcortical regions during the loss versus neutral condition (see Supplemental Table S1). These regions of interest were selected based on findings from previous meta-analyses [54–56].

*Genetic factors*
Genome-wide association studies (GWAS)' summary statistics allow for the estimation of a polygenic risk score in a genotyped independent sample. We leveraged previously published GWAS for four psychiatric diseases: major depression [57], bipolar disorder [58], anorexia [59] and schizophrenia [60]; as well as the most recent cross-disorder [61] GWAS performed by the psychiatric genomics consortium (PGC) (for methods see Supplemental Note 6).

**Supplemental Note 6: Polygenic risk scores method**

To avoid bias due to correlated SNPs arising from linkage-disequilibrium (LD), the GWAS summary statistics were subjected to a Bayesian analysis to approximate the results of a conditional GWAS (i.e. one estimating the effect for all SNPs simultaneously). This was done using the software SBayesR [62] implemented within a tool for Genome-wide Complex Trait Bayesian analysis (GCTB). The estimated conditional effect sizes were then used for polygenic scoring in the ABCD sample. The ABCD genotyping has been previously described [63]. Briefly, saliva samples were obtained at the baseline visit and genotyping was performed using a Smokescreen array following standard DNA extraction protocols. Quality control removed genetic variants with a low call rate (less than 99% of the sample), and samples with a missing rate greater than 20 percent or with conflicting IDs. This quality controlled dataset was then imputed to the 1000G Phase 3 reference panel using the Michigan Imputation Server[64]. Imputed genotype probabilities were extracted from the imputed data using QCTOOL v2. Only SNPs passing quality control (minor allele frequency >0.01, call rate > 0.9 and imputation score > 0.6) were included in the polygenic risk scores which were estimated using PLINKv2.

**Supplemental Table S1. Variables used in binomial penalized logistic regression analysis to predict group status (healthy controls vs. clinical controls vs. suicidal ideation or attempt)**

| Measure | Description | Assessment |
|---|---|---|
| **Sociodemographic** | | |
| Age | Chronological age in months | Demographics survey |
| Sex | Male or female | Demographics survey |
| Parental marital status | 1=married;<br>2=widowed;<br>3= divorced;<br>4=separated;<br>5=never married;<br>6=living with partner | Parent demographics survey |
| Family income in last 12 months | 1= Less than $5,000;<br>2=$5,000 through $11,999;<br>3=$12,000 through $15,999;<br>4=$16,000 through $24,999;<br>5=$25,000 through $34,999;<br>6=$35,000 through $49,999;<br>7=$50,000 through $74,999;<br>8= $75,000 through $99,999;<br>9=$100,000 through $199,999;<br>10=$200,000 and greater | Parent demographics survey |
| Parental education | 1=1st grade<br>2=2nd grade<br>3=3rd grade<br>4=4th grade<br>5=5th grade<br>6=6th grade<br>7=7th grade<br>8=8th grade<br>9=9th grade<br>10=10th grade<br>11=11th grade<br>12=12th grade<br>13=high School<br>14= GED or equivalent diploma<br>15=some college<br>16=associate degree: Occupational<br>17=associate degree: Academic program<br>18= bachelor's degree<br>19= master's degree<br>20= professional school degree<br>21=doctoral degree | Parent demographics survey: Highest grade or level of school or the highest degree completed: Calculated as parent education score + partner education score |
| Race of the child | Recoded to: white, black, native-american, asian, mixed,<br>other | Parent demographics survey |
| Number of cohabitants | Number of individuals living in the same household | Parent demographics survey |
| **Physical health** | | |
| Physical illness<br>(parent reported) | Dichotomous variable, scored 1 if parent reported that the child had a history of brain injury, cancer, cerebral palsy, diabetes, epilepsy, multiple sclerosis or heart disease | ABCD Medical History Questionnaire |
| Pubertal status<br>(child reported) | Categories:<br>1=prepuberty;<br>2=early puberty; | Pubertal Developmental Scale - categories based on sum scores |

| | | |
|---|---|---|
| | 3=mid puberty; 4=late puberty; 5=post puberty | |
| Sleep breathing disorders | Score on the sleep breathing disorder subscale | Sleep Disturbance Scale for Children |
| Sleep arousal disorders | Score on the sleep arousal disorder subscale | Sleep Disturbance Scale for Children |
| Sleep-wake transition disorders | Score on the sleep-wake transition disorder subscale | Sleep Disturbance Scale for Children |
| Sleep-excessive somnolence | Score on the excessive somnolence subscale | Sleep Disturbance Scale for Children |
| Sleep - hyperhydrosis | Score on the hyperhydrosis disorder subscale | Sleep Disturbance Scale for Children |
| Sleep disturbance - total score | Total score on the Sleep Disturbance Scale for Children | Sleep Disturbance Scale for Children |
| Physical activity in past week (child-reported) | Number of days in the past week during which the child was physically active for at least sixty minutes per day | Youth Risk Behavior Survey |
| Physical activity training muscles in past week (child-reported) | Number of days in the past week during which the child was did exercises to strengthen or tone muscles | Youth Risk Behavior Survey |
| Body Mass Index | Body mass index was calculated from height (in inches) and weight (in lbs) | Physical exam |
| Screen time weekday (child-reported) | Sum of hours during a weekday a child typically spends on a computer, cellphone, tablet or other electronic device | ABCD Youth Screen Time Survey |
| Screen time weekend (child-reported) | Sum of hours during a weekend day a child typically spends on a computer, cellphone, tablet or other electronic device | ABCD Youth Screen Time Survey |
| **Social environment** | | |
| Parental monitoring (youth-reported) | Responses to each item were coded as 1-5 and the mean of the responses was calculated | ABCD Parental Monitoring Survey |
| Family conflict (child-reported) | Responses to 9 items were coded 0/1 and a sum score was created | ABCD Family Environment Scale: Family Conflict Subscale Modified from PhenX |
| Prosocial behavior (child-reported) | Responses to 3 items were coded 0-2 and the mean was calculated | Prosocial Behavior Subscale from the Strengths and Difficulties Questionnaire |
| Parental acceptance (child-reported) | Responses to 5 items were coded 1-3 and the mean was calculated | Acceptance subscale from the Children's reported of Parental Behavioral Inventory |
| Bullying (child-reported) | Problems with bullying at school or in the neighborhood: coded yes/no | Introduction to the K-SADS5 |

| | | |
|---|---|---|
| Best friend (parent-reported) | Does the child have a best friend: coded yes/no/unsure | Introduction to the K-SADS5 |
| Friend group (parent-reported) | Does the child have a friend group: coded: yes/no/unsure | Introduction to the K-SADS5 |
| Neighborhood safety (parent reported) | Mean of three items | ABCD Neighborhood Safety/Crime Survey Modified from PhenX |
| School environment (child-reported) | Responses to 5 items were coded 1-4 and a sumscore was calculated | ABCD School Risk and Protective Factors Survey - Environment Subscale |
| School involvement (child-reported) | Responses to 4 items were coded 1-4 and a sumscore was created | ABCD School Risk and Protective Factors Survey - Involvement Subscale |
| School disengagement (child-reported) | Responses to 2 items were coded 1-4 and a sumscore was created | ABCD School Risk and Protective Factors Survey Disengagement Subscale |
| **Clinical psychiatric (only included in analyses with clinical controls & suicide group)** | | |
| CBCL Anxious Depression | T-score for the anxious/depressed subscale | Achenbach Child Behavior Checklist |
| CBCL Withdrawn Depressed | T-score for the withdrawn/depressed subscale | Achenbach Child Behavior Checklist |
| CBCL Somatic Complaints | T-score for the somatic complaints subscale | Achenbach Child Behavior Checklist |
| CBCL Social Problems | T-score for the social problems subscale | Achenbach Child Behavior Checklist |
| CBCL Thought Problems | T-score for the thought problems subscale | Achenbach Child Behavior Checklist |
| CBCL Attention Problems | T-score for the attention problems subscale | Achenbach Child Behavior Checklist |
| CBCL Rule Breaking behavior | T-score for the rule-breaking behavior subscale | Achenbach Child Behavior Checklist |
| CBCL Aggressive behavior | T-score for the aggressive behavior subscale | Achenbach Child Behavior Checklist |
| CBCL Internalizing Broad Band Score | T-score for the internalizing broad band score | Achenbach Child Behavior Checklist |
| CBCL Externalizing Broad Band Score | T-score for the externalizing broad band score | Achenbach Child Behavior Checklist |
| CBCL Total problems Score | T-score for the total problems score | Achenbach Child Behavior Checklist |
| CBCL Depression | T-score for the DSM5 depression subscale | Achenbach Child Behavior Checklist |
| CBCL Anxiety Disorder | T-score for the DSM5 anxiety disorder subscale | Achenbach Child Behavior Checklist |
| CBCL Somatic | T-score for the DSM5 somatic | Achenbach Child Behavior Checklist |

| | | |
|---|---|---|
| | complaints subscale | |
| CBCL ADHD | T-score for the DSM5 ADHD subscale | Achenbach Child Behavior Checklist |
| CBCL Oppositional Defiant Problems | T-score for the DSM5 ODD subscale | Achenbach Child Behavior Checklist |
| CBCL Conduct Problems | T-score for the DSM5 conduct problems subscale | Achenbach Child Behavior Checklist |
| CBCL Sluggish Cognitive Tempo | T-score for the sluggish cognitive tempo subscale | Achenbach Child Behavior Checklist |
| CBCL Obsessive-compulsive problems | T-score for the DSM5 OCD subscale | Achenbach Child Behavior Checklist |
| CBCL Stress | T-score for the stress tempo subscale | Achenbach Child Behavior Checklist |
| Prodromal psychosis | 21 items scored 1-5 and 1 item scored 0-1, a sum score is calculated | Brief version of the Pediatric Psychosis Questionnaire |
| Mania symptoms | 10 items scored 1-3, a sum score is calculated | Mania Scale from the Parent General Behavior Inventory |
| Mental health service use | Has the child ever received mental health or substance abuse services? Coded yes/no | Introduction to the K-SADS5 |
| Service use - Outpatient care | Has the child ever received outpatient mental health services? Coded yes/no | Introduction to the K-SADS5 |
| Service use - Partial hospitalization | Has the child ever been partially hospitalized for mental health issues? Coded yes/no | Introduction to the K-SADS5 |
| Service use - Inpatient care | Has the child ever received inpatient mental health services? Coded yes/no | Introduction to the K-SADS5 |
| Service use - Psychotherapy | Has the child ever received psychotherapy? Coded yes/no | Introduction to the K-SADS5 |
| Service use - Medication | Has the child ever been prescribed medication for mental health issues? Coded yes/no | Introduction to the K-SADS5 |
| Service use - Other treatment | Has the child ever received other types of treatment for mental health issues? Coded yes/no | Introduction to the K-SADS5 |
| Service use - Clinical treatment | Has the child not received treatment for mental health issues? Coded yes/no | Introduction to the K-SADS5 |
| Bipolar disorder - present | Current diagnosis of bipolar disorder. Coded yes/no | K-SADS5 (parent reported) |
| Depressive disorder - present | Current diagnosis of a depressive disorder. Coded yes/no | K-SADS5 (parent reported) |
| Psychotic disorder - present | Current diagnosis of a psychotic disorder. Coded yes/no | K-SADS5 (parent reported) |
| ADHD - present | Current diagnosis of ADHD disorder. Coded yes/no | K-SADS5 (parent reported) |

| | | |
|---|---|---|
| PD - present | Current diagnosis of panic disorder. Coded yes/no | K-SADS5 (parent reported) |
| SA - present | Current diagnosis of social anxiety. Coded yes/no | K-SADS5 (parent reported) |
| SP - present | Current diagnosis of specific phobia. Coded yes/no | K-SADS5 (parent reported) |
| Generalized anxiety disorder - present | Current diagnosis of generalized anxiety disorder. Coded yes/no | K-SADS5 (parent reported) |
| Substance use disorder - present | Current diagnosis of substance use disorder. Coded yes/no | K-SADS5 (parent reported) |
| Obsessive compulsive disorder - present | Current diagnosis of obsessive compulsive disorder. Coded yes/no | K-SADS5 (parent reported) |
| Post-traumatic stress disorder - present | Current diagnosis of post-traumatic stress disorder. Coded yes/no | K-SADS5 (parent reported) |
| Eating disorder - present | Current diagnosis of an eating disorder. Coded yes/no | K-SADS5 (parent reported) |
| Bipolar disorder - past | Past diagnosis of bipolar disorder. Coded yes/no | K-SADS5 (parent reported) |
| Depressive disorder - past | Past diagnosis of a depressive disorder. Coded yes/no | K-SADS5 (parent reported) |
| Psychotic disorder - past | Past diagnosis of a psychotic disorder. Coded yes/no | K-SADS5 (parent reported) |
| ADHD - past | Past diagnosis of ADHD disorder. Coded yes/no | K-SADS5 (parent reported) |
| PD - past | Past diagnosis of panic disorder. Coded yes/no | K-SADS5 (parent reported) |
| SA - past | Past diagnosis of social anxiety. Coded yes/no | K-SADS5 (parent reported) |
| SP - past | Past diagnosis of specific phobia. Coded yes/no | K-SADS5 (parent reported) |
| Generalized anxiety disorder - past | Past diagnosis of generalized anxiety disorder. Coded yes/no | K-SADS5 (parent reported) |
| Substance use disorder - past | Past diagnosis of substance use disorder. Coded yes/no | K-SADS5 (parent reported) |
| Eating disorder - past | Past diagnosis of an eating disorder. Coded yes/no | K-SADS5 (parent reported) |
| Depressive disorder (child-reported) | Lifetime diagnosis of a depressive disorder. Coded yes/no | K-SADS5 (child reported) |
| Bipolar disorder (child-reported) | Lifetime diagnosis of bipolar disorder. Coded yes/no | K-SADS5 (child reported) |
| Anxiety disorder (child-reported) | Lifetime diagnosis of an anxiety disorder. Coded yes/no | K-SADS5 (child reported) |
| Family history of psychopathology | Dichotomous variable, with 'yes' indicating a family history of mania, psychosis, nerves, mental health hospitalization or mental health treatment | Family History Assessment from NCANDA |
| **Cognition** | | |

| | | |
|---|---|---|
| Flanker inhibitory control | Age-corrected standard score | NIH Toolbox Flanker Inhibitory Control and Attention Test |
| Card Sorting | Age-corrected standard score | NIH Toolbox Dimensional Change Card Sort Test |
| Cash choice task | Would you rather have $75 in three days or $115 in 3 months? Scored 1-2 | Cash Choice Task - Delay Discounting |
| Impulsivity - negative urgency | 4 items scored 1-4 a sum score is created | UPPS-P Negative Urgency subscale |
| Impulsivity - lack of planning | 4 items scored 1-4 a sum score is created | UPPS-P Lack of Planning subscale |
| Impulsivity- sensation seeking | 4 items scored 1-4 a sum score is created | UPPS-P Sensation Seeking subscale |
| Impulsivity - positive urgency | 4 items scored 1-4 a sum score is created | UPPS-P Positive Urgency subscale |
| Impulsivity - lack of perseverance | 4 items scored 1-4 a sum score is created | UPPS-P Lack of Perseverance subscale |
| Picture vocabulary | Age-corrected standard score | NIH Toolbox Picture Vocabulary Test |
| List sorting task | Age-corrected standard score | NIH Toolbox List Sorting Working Memory Test |
| Pattern comparison | Age-corrected standard score | NIH Toolbox Pattern Comparison Processing Speed Test |
| Picture sequence memory | Age-corrected standard score | NIH Toolbox Picture Sequence Memory Test |
| Reading test | Age-corrected standard score | NIH Toolbox Oral Reading Recognition Test |
| Matrix reasoning | Total scaled score | WISC-V Matrix Reasoning |
| **Neuroimaging** | | |
| Correct stop vs correct go - left thalamus | Mean beta weight for SST correct stop versus correct go contrast | Stop Signal task - Correct stop vs. correct go contrast |
| Correct stop vs correct go - left caudate | Mean beta weight for SST correct stop versus correct go contrast | Stop Signal task - Correct stop vs. correct go contrast |
| Correct stop vs correct go - left putamen | Mean beta weight for SST correct stop versus correct go contrast | Stop Signal task - Correct stop vs. correct go contrast |
| Correct stop vs correct go - left pallidum | Mean beta weight for SST correct stop versus correct go contrast | Stop Signal task - Correct stop vs. correct go contrast |
| Correct stop vs correct go - right thalamus | Mean beta weight for SST correct stop versus correct go contrast | Stop Signal task - Correct stop vs. correct go contrast |
| Correct stop vs correct go - right caudate | Mean beta weight for SST correct stop versus correct go contrast | Stop Signal task - Correct stop vs. correct go contrast |
| Correct stop vs correct go - right putamen | Mean beta weight for SST correct stop versus correct go contrast | Stop Signal task - Correct stop vs. correct go contrast |
| Correct stop vs correct go - right pallidum | Mean beta weight for SST correct stop versus correct go contrast | Stop Signal task - Correct stop vs. correct go contrast |
| Correct stop vs correct go - left inferior parietal gyrus | Mean beta weight for SST correct stop versus correct go contrast | Stop Signal task - Correct stop vs. correct go contrast |

| | | |
|---|---|---|
| Correct stop vs correct go - left pars opercularis | Mean beta weight for SST correct stop versus correct go contrast | Stop Signal task - Correct stop vs. correct go contrast |
| Correct stop vs correct go - left pars oribitalis | Mean beta weight for SST correct stop versus correct go contrast | Stop Signal task - Correct stop vs. correct go contrast |
| Correct stop vs correct go - left triangularis | Mean beta weight for SST correct stop versus correct go contrast | Stop Signal task - Correct stop vs. correct go contrast |
| Correct stop vs correct go - left precentral | Mean beta weight for SST correct stop versus correct go contrast | Stop Signal task - Correct stop vs. correct go contrast |
| Correct stop vs correct go - left superior temporal | Mean beta weight for SST correct stop versus correct go contrast | Stop Signal task - Correct stop vs. correct go contrast |
| Correct stop vs correct go - left insula | Mean beta weight for SST correct stop versus correct go contrast | Stop Signal task - Correct stop vs. correct go contrast |
| Correct stop vs correct go - right inferior parietal gyrus | Mean beta weight for SST correct stop versus correct go contrast | Stop Signal task - Correct stop vs. correct go contrast |
| Correct stop vs correct go - right pars opercularis | Mean beta weight for SST correct stop versus correct go contrast | Stop Signal task - Correct stop vs. correct go contrast |
| Correct stop vs correct go - right pars orbitalis | Mean beta weight for SST correct stop versus correct go contrast | Stop Signal task - Correct stop vs. correct go contrast |
| Correct stop vs correct go - right triangularis | Mean beta weight for SST correct stop versus correct go contrast | Stop Signal task - Correct stop vs. correct go contrast |
| Correct stop vs correct go - right precentral gyrus | Mean beta weight for SST correct stop versus correct go contrast | Stop Signal task - Correct stop vs. correct go contrast |
| Correct stop vs correct go - right superior temporal gyrus | Mean beta weight for SST correct stop versus correct go contrast | Stop Signal task - Correct stop vs. correct go contrast |
| Correct stop vs correct go - right insula | Mean beta weight for SST correct stop versus correct go contrast | Stop Signal task - Correct stop vs. correct go contrast |
| Reward vs neutral - left thalamus | Mean beta weight for MID anticipation of reward versus neutral contrast | Monetary Incentive Delay task |
| Reward vs neutral - left caudate | Mean beta weight for MID anticipation of reward versus neutral contrast | Monetary Incentive Delay task |
| Reward vs neutral - left putamen | Mean beta weight for MID anticipation of reward versus neutral contrast | Monetary Incentive Delay task |
| Reward vs neutral - left pallidum | Mean beta weight for MID anticipation of reward versus neutral contrast | Monetary Incentive Delay task |
| Reward vs neutral - left amygdala | Mean beta weight for MID anticipation of reward versus neutral contrast | Monetary Incentive Delay task |
| Reward vs neutral - left accumbens | Mean beta weight for MID anticipation of reward versus neutral contrast | Monetary Incentive Delay task |
| Reward vs neutral - right thalamus | Mean beta weight for MID anticipation of reward versus neutral contrast | Monetary Incentive Delay task |
| Reward vs neutral - right caudate | Mean beta weight for MID anticipation of reward versus neutral contrast | Monetary Incentive Delay task |

| | | |
|---|---|---|
| Reward vs neutral - right putamen | Mean beta weight for MID anticipation of reward versus neutral contrast | Monetary Incentive Delay task |
| Reward vs neutral - right pallidum | Mean beta weight for MID anticipation of reward versus neutral contrast | Monetary Incentive Delay task |
| Reward vs neutral - right amygdala | Mean beta weight for MID anticipation of reward versus neutral contrast | Monetary Incentive Delay task |
| Reward vs neutral - right accumbens | Mean beta weight for MID anticipation of reward versus neutral contrast | Monetary Incentive Delay task |
| Reward vs neutral - left caudal middle frontal gyrus | Mean beta weight for MID anticipation of reward versus neutral contrast | Monetary Incentive Delay task |
| Reward vs neutral - left cuneus | Mean beta weight for MID anticipation of reward versus neutral contrast | Monetary Incentive Delay task |
| Reward vs neutral - left inferior temporal gyrus | Mean beta weight for MID anticipation of reward versus neutral contrast | Monetary Incentive Delay task |
| Reward vs neutral - left parahippocampal gyrus | Mean beta weight for MID anticipation of reward versus neutral contrast | Monetary Incentive Delay task |
| Reward vs neutral - left paracentral gyrus | Mean beta weight for MID anticipation of reward versus neutral contrast | Monetary Incentive Delay task |
| Reward vs neutral - left precentral gyrus | Mean beta weight for MID anticipation of reward versus neutral contrast | Monetary Incentive Delay task |
| Reward vs neutral - left rostral middle frontal gyrus | Mean beta weight for MID anticipation of reward versus neutral contrast | Monetary Incentive Delay task |
| Reward vs neutral - left superior frontal gyrus | Mean beta weight for MID anticipation of reward versus neutral contrast | Monetary Incentive Delay task |
| Reward vs neutral - left superior temporal gyrus | Mean beta weight for MID anticipation of reward versus neutral contrast | Monetary Incentive Delay task |
| Reward vs neutral - left supramarginal gyrus | Mean beta weight for MID anticipation of reward versus neutral contrast | Monetary Incentive Delay task |
| Reward vs neutral - left insula | Mean beta weight for MID anticipation of reward versus neutral contrast | Monetary Incentive Delay task |
| Reward vs neutral - right caudal middle frontal gyrus | Mean beta weight for MID anticipation of reward versus neutral contrast | Monetary Incentive Delay task |
| Reward vs neutral - right cuneus | Mean beta weight for MID anticipation of reward versus neutral contrast | Monetary Incentive Delay task |
| Reward vs neutral - right inferior temporal gyrus | Mean beta weight for MID anticipation of reward versus neutral contrast | Monetary Incentive Delay task |

| | | |
|---|---|---|
| Reward vs neutral - right parahippocampal gyrus | Mean beta weight for MID anticipation of reward versus neutral contrast | Monetary Incentive Delay task |
| Reward vs neutral - right paracentral gyrus | Mean beta weight for MID anticipation of reward versus neutral contrast | Monetary Incentive Delay task |
| Reward vs neutral - right precentral gyrus | Mean beta weight for MID anticipation of reward versus neutral contrast | Monetary Incentive Delay task |
| Reward vs neutral - right rostral middle frontal gyrus | Mean beta weight for MID anticipation of reward versus neutral contrast | Monetary Incentive Delay task |
| Reward vs neutral - right superior frontal gyrus | Mean beta weight for MID anticipation of reward versus neutral contrast | Monetary Incentive Delay task |
| Reward vs neutral - right superior temporal gyrus | Mean beta weight for MID anticipation of reward versus neutral contrast | Monetary Incentive Delay task |
| Reward vs neutral - right supramarginal gyrus | Mean beta weight for MID anticipation of reward versus neutral contrast | Monetary Incentive Delay task |
| Reward vs neutral - right insula | Mean beta weight for MID anticipation of reward versus neutral contrast | Monetary Incentive Delay task |
| Loss vs neutral - left thalamus | Mean beta weight for MID anticipation of loss versus neutral contrast | Monetary Incentive Delay task |
| Loss vs neutral - left caudate | Mean beta weight for MID anticipation of loss versus neutral contrast | Monetary Incentive Delay task |
| Loss vs neutral - left putamen | Mean beta weight for MID anticipation of loss versus neutral contrast | Monetary Incentive Delay task |
| Loss vs neutral - left pallidum | Mean beta weight for MID anticipation of loss versus neutral contrast | Monetary Incentive Delay task |
| Loss vs neutral - left hippocampus | Mean beta weight for MID anticipation of loss versus neutral contrast | Monetary Incentive Delay task |
| Loss vs neutral - left amygdala | Mean beta weight for MID anticipation of loss versus neutral contrast | Monetary Incentive Delay task |
| Loss vs neutral - left accumbens | Mean beta weight for MID anticipation of loss versus neutral contrast | Monetary Incentive Delay task |
| Loss vs neutral - right thalamus | Mean beta weight for MID anticipation of loss versus neutral contrast | Monetary Incentive Delay task |
| Loss vs neutral - right caudate | Mean beta weight for MID anticipation of loss versus neutral contrast | Monetary Incentive Delay task |
| Loss vs neutral - right putamen | Mean beta weight for MID anticipation of loss versus neutral contrast | Monetary Incentive Delay task |

| | | |
|---|---|---|
| Loss vs neutral - right pallidum | Mean beta weight for MID anticipation of loss versus neutral contrast | Monetary Incentive Delay task |
| Loss vs neutral - right hippocampus | Mean beta weight for MID anticipation of loss versus neutral contrast | Monetary Incentive Delay task |
| Loss vs neutral - right amygdala | Mean beta weight for MID anticipation of loss versus neutral contrast | Monetary Incentive Delay task |
| Loss vs neutral - right accumbens | Mean beta weight for MID anticipation of loss versus neutral contrast | Monetary Incentive Delay task |
| Loss vs neutral - left caudal anterior cingulate cortex | Mean beta weight for MID anticipation of loss versus neutral contrast | Monetary Incentive Delay task |
| Loss vs neutral - left caudal middle frontal gyrus | Mean beta weight for MID anticipation of loss versus neutral contrast | Monetary Incentive Delay task |
| Loss vs neutral - left fusiform gyrus | Mean beta weight for MID anticipation of loss versus neutral contrast | Monetary Incentive Delay task |
| Loss vs neutral - left precentral gyrus | Mean beta weight for MID anticipation of loss versus neutral contrast | Monetary Incentive Delay task |
| Loss vs neutral - left rostral anterior cingulate cortex | Mean beta weight for MID anticipation of loss versus neutral contrast | Monetary Incentive Delay task |
| Loss vs neutral - left rostral middle frontal gyrus | Mean beta weight for MID anticipation of loss versus neutral contrast | Monetary Incentive Delay task |
| Loss vs neutral - left superior frontal gyrus | Mean beta weight for MID anticipation of loss versus neutral contrast | Monetary Incentive Delay task |
| Loss vs neutral - left supramarginal gyrus | Mean beta weight for MID anticipation of loss versus neutral contrast | Monetary Incentive Delay task |
| Loss vs neutral - left insula | Mean beta weight for MID anticipation of loss versus neutral contrast | Monetary Incentive Delay task |
| Loss vs neutral - right caudal anterior cingulate cortex | Mean beta weight for MID anticipation of loss versus neutral contrast | Monetary Incentive Delay task |
| Loss vs neutral - right caudal middle frontal gyrus | Mean beta weight for MID anticipation of loss versus neutral contrast | Monetary Incentive Delay task |
| Loss vs neutral - right fusiform gyrus | Mean beta weight for MID anticipation of loss versus neutral contrast | Monetary Incentive Delay task |
| Loss vs neutral - right precentral gyrus | Mean beta weight for MID anticipation of loss versus neutral contrast | Monetary Incentive Delay task |
| Loss vs neutral - right rostral anterior cingulate cortex | Mean beta weight for MID anticipation of loss versus neutral contrast | Monetary Incentive Delay task |

| | | |
|---|---|---|
| Loss vs neutral - right rostral middle frontal gyrus | Mean beta weight for MID anticipation of loss versus neutral contrast | Monetary Incentive Delay task |
| Loss vs neutral - right superior frontal gyrus | Mean beta weight for MID anticipation of loss versus neutral contrast | Monetary Incentive Delay task |
| Loss vs neutral - right supramarginal gyrus | Mean beta weight for MID anticipation of loss versus neutral contrast | Monetary Incentive Delay task |
| Loss vs neutral - right insula | Mean beta weight for MID anticipation of loss versus neutral contrast | Monetary Incentive Delay task |
| **Genetic** | | |
| Depression polygenic risk score | Weighted sum of allele SBayesR conditional GWAS effect sizes. | Genotyping |
| Bipolar disorder polygenic risk score | Weighted sum of allele SBayesR conditional GWAS effect sizes. | Genotyping |
| Schizophrenia polygenic risk score | Weighted sum of allele SBayesR conditional GWAS effect sizes. | Genotyping |
| Anorexia polygenic risk score | Weighted sum of allele SBayesR conditional GWAS effect sizes. | Genotyping |
| Cross-disorder (psychiatric) polygenic risk score | Weighted sum of allele SBayesR conditional GWAS effect sizes. | Genotyping |

### Supplemental Table S2. Demographic in the train and test set (in child-reported and parent-reported groups)

| **Child reported** | Training set | | | Validation set | | |
|---|---|---|---|---|---|---|
| | HC (N=1650) | CC (N=2122) | STB (N=396) | HC (N=717) | CC (N=863) | STB (N=132) |
| Age | 9.9 | 9.9 | 9.9 | 9.9 | 10.0 | 9.9 |
| Sex (%F, n) | 51.5% (849) | 48.0% (1017) | 44.2% (175) | 56.1% (402) | 46.6% (402) | 47.7% (63) |
| Ideation (%, n) | NA | NA | 86.9% (344) | NA | NA | 88.6% (117) |
| Attempt (%, n) | NA | NA | 13.1% (52) | NA | NA | 11.3% (15) |
| **Parent reported** | Training set | | | Validation set | | |
| | HC (N=1683) | CC (N=2128) | STB (N=361) | HC (N=732) | CC (N=848) | STB (N=133) |
| Age | 9.9 | 9.9 | 9.9 | 10.0 | 10.0 | 10.0 |
| Sex (%F, n) | 51.8% (871) | 48.5% (1032) | 38.8% (140) | 55.8% (409) | 48.0% (407) | 37.6% (50) |
| Ideation (%, n) | NA | NA | 94.7% (342) | NA | NA | 91.7% (122) |
| Attempt (%, n) | NA | NA | 5.3% (19) | NA | NA | 8.3% (11) |

Note: CC: clinical control group; HC: healthy control group; STB: suicidal thoughts and behaviors group

**Supplemental Table S3. Performance measures of group classification in the overlapping sample (consistent parent and child-reported STB)**

| Comparison | Alpha | AUROC | SD AUROC | Sensitivity | Specificity | Accuracy |
|---|---|---|---|---|---|---|
| HC vs. CC | | | | | | |
| | 0.25 | 0.689 | 0.021 | 0.685 | 0.589 | 0.637 |
| | 0.50 | 0.689 | 0.023 | 0.688 | 0.587 | 0.637 |
| | 0.75 | 0.691 | 0.022 | 0.686 | 0.589 | 0.638 |
| | 1.00 | 0.688 | 0.021 | 0.688 | 0.586 | 0.637 |
| HC vs. STB | | | | | | |
| | 0.25 | 0.878 | 0.056 | 0.764 | 0.825 | 0.795 |
| | 0.50 | 0.879 | 0.064 | 0.766 | 0.828 | 0.797 |
| | 0.75 | 0.876 | 0.063 | 0.749 | 0.832 | 0.791 |
| | 1.00 | 0.886 | 0.055 | 0.751 | 0.839 | 0.795 |
| CC vs. STB | | | | | | |
| | 0.25 | 0.860 | 0.069 | 0.764 | 0.804 | 0.784 |
| | 0.50 | 0.865 | 0.070 | 0.761 | 0.822 | 0.792 |
| | 0.75 | 0.849 | 0.069 | 0.747 | 0.795 | 0.771 |
| | 1.00 | 0.858 | 0.067 | 0.743 | 0.806 | 0.774 |

Note: CC: clinical control group; HC: healthy control group; STB: suicidal thoughts and behaviors group

### Supplemental Table S4. Results of feature selection (child-reported)

| Comparison | Alpha: | 0.25 | 0.5 | 0.75 | 1 |
|---|---|---|---|---|---|
| HC vs. CC | | Age | Age | Age | Age |
| | | Race | Parental income | Parental income | Parental income |
| | | Parental income | Sleep: excessive somnolesence | Sleep: total score | Sleep: total score |
| | | Sleep: excessive somnolesence | Sleep: total score | BMI | BMI |
| | | Sleep: total score | BMI | Family conflict | Family conflict |
| | | BMI | Screen time weekend | School involvement | School involvement |
| | | Exercise in the past week | Family conflict | Bullying | Bullying |
| | | Screen time weekend | School involvement | Family history of psychopathology | Family history of psychopathology |
| | | Family conflict | Bullying | Impulsivity: positive urgency | Impulsivity: positive urgency |
| | | School involvement | Family history of psychopathology | Impulsivity: lack of perseverance | Impulsivity: lack of perseverance |
| | | Bullying | Impulsivity: positive urgency | Picture sequence memory task performance | Picture sequence memory task performance |
| | | Family history of psychopathology | Impulsivity: lack of perseverance | Left caudate activation during loss anticipation | Left caudate activation during loss anticipation |
| | | Cash Choice Task performance | Picture sequence memory task performance | Right inferior temporal lobe activation during reward anticipation | |
| | | Impulsivity: sensation seeking | Left caudate activation during loss anticipation | | |
| | | Impulsivity: positive urgency | | | |
| | | Impulsivity: lack of perseverance | | | |
| | | Picture sequence memory task performance | | | |
| | | Right inferior temporal lobe activation during reward anticipation | | | |
| | | Left caudate activation during loss anticipation | | | |
| HC vs. STB | | Sleep: excessive somnolesence | Sleep: total score | Sleep: total score | Sleep: total score |
| | | Sleep: total score | screen time weekend | screen time weekend | screen time weekend |
| | | screen time weekend | Parental monitoring | Parental monitoring | Family conflict |
| | | Parental monitoring | Family conflict | Family conflict | Bullying |
| | | Family conflict | School involvement | School involvement | Impulsivity: negative |

| | | | | |
|---|---|---|---|---|
| | | | | urgency |
| | School involvement | Pubertal status | Bullying | Impulsivity: lack of planning |
| | Pubertal status | Bullying | Impulsivity: negative urgency | |
| | Bullying | Impulsivity: negative urgency | Impulsivity: lack of planning | |
| | Having a group of friends | Impulsivity: lack of planning | | |
| | Impulsivity: negative urgency | | | |
| | Impulsivity: lack of perseverance | | | |
| | Impulsivity: lack of planning | | | |
| CC vs. STB | Race | Family conflict | Family conflict | Family conflict |
| | Family conflict | Prodromal psychosis | Prodromal psychosis | Prodromal psychosis |
| | Prodromal psychosis | CBCL: DSM5 Depression subscale | Impulsivity: negative urgency | Impulsivity: negative urgency |
| | CBCL: DSM5 Depression subscale | Impulsivity: negative urgency | Impulsivity: lack of planning | Impulsivity: lack of planning |
| | Impulsivity: negative urgency | Impulsivity: lack of planning | | |
| | Impulsivity: lack of planning | | | |

Note: CC: clinical control group; HC: healthy control group; STB: suicidal thoughts and behaviors group

### Supplemental Table S5. Results of feature selection (parent-reported)

| Comparison | Alpha: | 0.25 | 0.5 | 0.75 | 1 |
|---|---|---|---|---|---|
| HC vs. CC | | Age | Age | Age | Age |
| | | Race | Race | Race | Race |
| | | Parental income | Parental income | Parental income | Parental income |
| | | Sleep-excessive somnolesence | Sleep - total sleep score | Sleep - total sleep score | Sleep - total sleep score |
| | | Sleep - total sleep score | Exercise in the past week | BMI | BMI |
| | | Exercise in the past week | BMI | Screen time in weekends | Screen time in weekends |
| | | BMI | Screen time in weekends | Family conflict | Family conflict |
| | | Pubertal status | Family conflict | School involvement | School involvement |
| | | Screen time in weekends | School involvement | Bullying | Bullying |
| | | Family conflict | Bullying | Friend group | Family history of psychopathology |
| | | Neighborhood safety | Friend group | Family history of psychopathology | Impulsivity: sensation seeking |
| | | School involvement | Family history of psychopathology | Impulsivity: sensation seeking | Impulsivity: positive urgency |
| | | Bullying | Impulsivity: sensation seeking | Impulsivity: positive urgency | Impulsivity: lack of perseverance |
| | | Friend group | Impulsivity: positive urgency | Impulsivity: lack of perseverance | Impulsivity: negative urgency |
| | | Family history of psychopathology | Impulsivity: lack of perseverance | Impulsivity: negative urgency | Impulsivity: lack of planning |
| | | Cash Choice Task performance | Impulsivity: negative urgency | Impulsivity: lack of planning | Picture sequence memory |
| | | Impulsivity: sensation seeking | Impulsivity: lack of planning | Picture sequence memory | Pattern comparison: processing speed |
| | | Impulsivity: positive urgency | Pattern comparison: processing speed | Left caudate activation during loss anticipation | Left caudate activation during loss anticipation |
| | | Impulsivity: lack of perseverance | Picture sequence memory | Right amygdala activation during loss anticipation | Right amygdala activation during loss anticipation |
| | | Impulsivity: negative urgency | Left caudate activation during loss anticipation | | |
| | | Impulsivity: lack of planning | Right amygdala activation during loss anticipation | | |
| | | Pattern comparison: processing speed | | | |
| | | Picture sequence memory | | | |

| | | | | |
|---|---|---|---|---|
| | Left caudate activation during loss anticipation<br><br>Right amygdala activation during loss anticipation<br><br>Right thalamus activation during successful response inhibition<br><br>Right inferior temporal gyrus during reward anticipation | | | |
| HC vs. STB | Sex<br><br>Sleep-excessive somnolesence<br><br>Sleep - total sleep score<br><br>Family conflict<br><br>School environment<br><br>Bullying<br><br>Family history of psychopathology<br><br>Impulsivity: negative urgency<br><br>Impulsivity: lack of perseverance | Sex<br><br>Sleep-excessive somnolesence<br><br>Sleep - total sleep score<br><br>Family conflict<br><br>Bullying<br><br>Family history of psychopathology<br><br>Impulsivity: negative urgency | Sleep - total sleep score<br><br>Family conflict<br><br>Bullying<br><br>Family history of psychopathology<br><br>Impulsivity: negative urgency | Sleep - total sleep score<br><br>Family conflict<br><br>Bullying<br><br>Family history of psychopathology<br><br>Impulsivity: negative urgency |
| CC vs. STB | CBCL: Anxious Depression subscale<br><br>CBCL: Internalizing Broad Band Score<br><br>CBCL: Externalizing Broad Band Score<br><br>CBCL: DSM5 Depression subscale<br><br>CBCL: Conduct Disorder subscale<br><br>History of mental health service use<br><br>History of mental health treatment | CBCL: Anxious Depression subscale<br><br>CBCL: Internalizing Broad Band Score<br><br>CBCL: Externalizing Broad Band Score<br><br>CBCL: DSM5 Depression subscale<br><br>History of mental health treatment | CBCL: Anxious Depression subscale<br><br>CBCL: Internalizing Broad Band Score<br><br>CBCL: Externalizing Broad Band Score<br><br>CBCL: DSM5 Depression subscale<br><br>History of mental health treatment | CBCL: Internalizing Broad Band Score<br><br>CBCL: Externalizing Broad Band Score<br><br>CBCL: DSM5 Depression subscale<br><br>History of mental health treatment |

Note: CC: clinical control group;  HC: healthy control group; STB: suicidal thoughts and behaviors group

**Supplemental Table S6. Classification of groups (child-reported) in independent dataset using only the features selected at different alpha levels in the training dataset**

| Comparison | Using features selected in training dataset at alpha | AUROC | Sensitivity | Specificity | Accuracy |
|---|---|---|---|---|---|
| HC vs. CC | | | | | |
| | 0.25 | 0.704 | 0.660 | 0.633 | 0.646 |
| | 0.50 | 0.701 | 0.665 | 0.634 | 0.650 |
| | 0.75 | 0.701 | 0.656 | 0.633 | 0.644 |
| | 1.00 | 0.701 | 0.661 | 0.638 | 0.650 |
| HC vs. STB | | | | | |
| | 0.25 | 0.790 | 0.735 | 0.649 | 0.692 |
| | 0.50 | 0.786 | 0.727 | 0.649 | 0.688 |
| | 0.75 | 0.792 | 0.758 | 0.664 | 0.711 |
| | 1.00 | 0.784 | 0.720 | 0.657 | 0.688 |
| CC vs. STB | | | | | |
| | 0.25 | 0.683 | 0.697 | 0.568 | 0.632 |
| | 0.50 | 0.703 | 0.712 | 0.567 | 0.639 |
| | 0.75 | 0.711 | 0.758 | 0.553 | 0.655 |
| | 1.00 | 0.711 | 0.758 | 0.553 | 0.655 |

Note: CC: clinical control group; HC: healthy control group; STB: suicidal thoughts and behaviors group

**Supplemental Table S7. Classification of groups (parent-reported) in independent dataset using only the features selected at different alpha levels in the training dataset**

| Comparison | Using features selected in training dataset at alpha | AUROC | Sensitivity | Specificity | Accuracy |
|---|---|---|---|---|---|
| HC vs. CC | | | | | |
| | 0.25 | 0.706 | 0.634 | 0.664 | 0.649 |
| | 0.50 | 0.707 | 0.627 | 0.669 | 0.648 |
| | 0.75 | 0.705 | 0.638 | 0.651 | 0.644 |
| | 1.00 | 0.703 | 0.635 | 0.647 | 0.641 |
| HC vs. STB | | | | | |
| | 0.25 | 0.817 | 0.752 | 0.721 | 0.737 |
| | 0.50 | 0.816 | 0.767 | 0.736 | 0.752 |
| | 0.75 | 0.807 | 0.752 | 0.724 | 0.738 |
| | 1.00 | 0.807 | 0.752 | 0.724 | 0.738 |
| CC vs. STB | | | | | |
| | 0.25 | 0.706 | 0.583 | 0.714 | 0.649 |
| | 0.50 | 0.706 | 0.583 | 0.700 | 0.642 |
| | 0.75 | 0.706 | 0.583 | 0.699 | 0.641 |
| | 1.00 | 0.703 | 0.598 | 0.700 | 0.649 |

Note: CC: clinical control group; HC: healthy control group; STB: suicidal thoughts and behaviors group

**Supplemental Table S8. Classification of STB groups (child-reported): Results of binomial penalized logistic regression analysis only including specific types of predictors**

| **Sociodemographic** | | | | | | |
|---|---|---|---|---|---|---|
| Comparison | Alpha | AUROC | SD AUROC | Sensitivity | Specificity | Accuracy |
| HC vs. CC | | | | | | |
| | 0.25 | 0.559 | 0.025 | 0.550 | 0.533 | 0.541 |
| | 0.50 | 0.558 | 0.030 | 0.539 | 0.537 | 0.538 |
| | 0.75 | 0.560 | 0.025 | 0.534 | 0.545 | 0.540 |
| | 1.00 | 0.557 | 0.029 | 0.540 | 0.541 | 0.541 |
| HC vs. STB | | | | | | |
| | 0.25 | 0.587 | 0.054 | 0.523 | 0.599 | 0.561 |
| | 0.50 | 0.588 | 0.066 | 0.523 | 0.617 | 0.570 |
| | 0.75 | 0.590 | 0.060 | 0.521 | 0.613 | 0.567 |
| | 1.00 | 0.585 | 0.063 | 0.526 | 0.601 | 0.563 |
| CC vs. STB | | | | | | |
| | 0.25 | 0.556 | 0.058 | 0.585 | 0.494 | 0.540 |
| | 0.50 | 0.552 | 0.060 | 0.592 | 0.479 | 0.536 |
| | 0.75 | 0.547 | 0.069 | 0.590 | 0.482 | 0.536 |
| | 1.00 | 0.540 | 0.055 | 0.644 | 0.419 | 0.532 |

| **Physical health** | | | | | | |
|---|---|---|---|---|---|---|
| Comparison | Alpha | AUROC | SD AUROC | Sensitivity | Specificity | Accuracy |
| HC vs. CC | | | | | | |
| | 0.25 | 0.668 | 0.021 | 0.690 | 0.550 | 0.620 |
| | 0.50 | 0.667 | 0.028 | 0.695 | 0.547 | 0.621 |
| | 0.75 | 0.667 | 0.028 | 0.696 | 0.550 | 0.623 |
| | 1.00 | 0.669 | 0.029 | 0.697 | 0.551 | 0.624 |
| HC vs. STB | | | | | | |
| | 0.25 | 0.712 | 0.052 | 0.596 | 0.717 | 0.657 |
| | 0.50 | 0.726 | 0.060 | 0.607 | 0.734 | 0.670 |
| | 0.75 | 0.717 | 0.056 | 0.601 | 0.728 | 0.664 |
| | 1.00 | 0.715 | 0.061 | 0.603 | 0.720 | 0.661 |
| CC vs. STB | | | | | | |
| | 0.25 | 0.583 | 0.063 | 0.544 | 0.578 | 0.561 |
| | 0.50 | 0.582 | 0.062 | 0.566 | 0.559 | 0.563 |
| | 0.75 | 0.579 | 0.062 | 0.540 | 0.566 | 0.553 |

| | | | | | | |
|---|---|---|---|---|---|---|
| | 1.00 | 0.582 | 0.057 | 0.547 | 0.570 | 0.559 |
| **Social environment** | | | | | | |
| Comparison | Alpha | AUROC | SD AUROC | Sensitivity | Specificity | Accuracy |
| HC vs. CC | | | | | | |
| | 0.25 | 0.604 | 0.024 | 0.694 | 0.458 | 0.576 |
| | 0.50 | 0.605 | 0.028 | 0.689 | 0.458 | 0.574 |
| | 0.75 | 0.604 | 0.031 | 0.694 | 0.459 | 0.576 |
| | 1.00 | 0.601 | 0.030 | 0.691 | 0.454 | 0.572 |
| HC vs. STB | | | | | | |
| | 0.25 | 0.738 | 0.738 | 0.618 | 0.741 | 0.680 |
| | 0.50 | 0.734 | 0.734 | 0.615 | 0.737 | 0.676 |
| | 0.75 | 0.737 | 0.737 | 0.618 | 0.748 | 0.683 |
| | 1.00 | 0.735 | 0.735 | 0.609 | 0.740 | 0.674 |
| CC vs. STB | | | | | | |
| | 0.25 | 0.633 | 0.062 | 0.553 | 0.645 | 0.599 |
| | 0.50 | 0.641 | 0.063 | 0.553 | 0.661 | 0.607 |
| | 0.75 | 0.639 | 0.056 | 0.548 | 0.658 | 0.603 |
| | 1.00 | 0.632 | 0.059 | 0.551 | 0.653 | 0.602 |
| **Clinical psychiatric** | | | | | | |
| Comparison | Alpha | AUROC | SD AUROC | Sensitivity | Specificity | Accuracy |
| CC vs. STB | | | | | | |
| | 0.25 | 0.676 | 0.059 | 0.532 | 0.721 | 0.627 |
| | 0.50 | 0.681 | 0.059 | 0.525 | 0.729 | 0.627 |
| | 0.75 | 0.676 | 0.059 | 0.523 | 0.719 | 0.621 |
| | 1.00 | 0.677 | 0.057 | 0.522 | 0.727 | 0.625 |
| **Cognition** | | | | | | |
| Comparison | Alpha | AUROC | SD AUROC | Sensitivity | Specificity | Accuracy |
| HC vs. CC | | | | | | |
| | 0.25 | 0.587 | 0.027 | 0.577 | 0.547 | 0.562 |
| | 0.50 | 0.587 | 0.022 | 0.580 | 0.547 | 0.564 |
| | 0.75 | 0.588 | 0.030 | 0.578 | 0.548 | 0.563 |
| | 1.00 | 0.587 | 0.029 | 0.579 | 0.545 | 0.562 |
| HC vs. STB | | | | | | |
| | 0.25 | 0.712 | 0.057 | 0.647 | 0.665 | 0.656 |
| | 0.50 | 0.712 | 0.050 | 0.642 | 0.670 | 0.656 |

| | | | | | | |
|---|---|---|---|---|---|---|
| | 0.75 | 0.712 | 0.056 | 0.644 | 0.669 | 0.656 |
| | 1.00 | 0.712 | 0.047 | 0.644 | 0.668 | 0.656 |
| CC vs. STB | | | | | | |
| | 0.25 | 0.645 | 0.060 | 0.599 | 0.608 | 0.604 |
| | 0.50 | 0.654 | 0.063 | 0.598 | 0.628 | 0.613 |
| | 0.75 | 0.648 | 0.067 | 0.599 | 0.613 | 0.606 |
| | 1.00 | 0.650 | 0.070 | 0.595 | 0.615 | 0.605 |

**Neuroimaging**

| Comparison | Alpha | AUROC | SD AUROC | Sensitivity | Specificity | Accuracy |
|---|---|---|---|---|---|---|
| HC vs. CC | | | | | | |
| | 0.25 | 0.503 | 0.020 | 0.583 | 0.421 | 0.502 |
| | 0.50 | 0.502 | 0.021 | 0.519 | 0.479 | 0.499 |
| | 0.75 | 0.503 | 0.021 | 0.581 | 0.423 | 0.502 |
| | 1.00 | 0.501 | 0.017 | 0.576 | 0.425 | 0.501 |
| HC vs. STB | | | | | | |
| | 0.25 | 0.535 | 0.053 | 0.576 | 0.481 | 0.529 |
| | 0.50 | 0.534 | 0.056 | 0.639 | 0.406 | 0.523 |
| | 0.75 | 0.534 | 0.055 | 0.603 | 0.439 | 0.521 |
| | 1.00 | 0.535 | 0.055 | 0.600 | 0.451 | 0.525 |
| CC vs. STB | | | | | | |
| | 0.25 | 0.496 | 0.025 | 0.758 | 0.241 | 0.500 |
| | 0.50 | 0.496 | 0.033 | 0.771 | 0.221 | 0.496 |
| | 0.75 | 0.504 | 0.031 | 0.792 | 0.212 | 0.502 |
| | 1.00 | 0.497 | 0.031 | 0.773 | 0.222 | 0.497 |

**Genetic**

| Comparison | Alpha | AUROC | SD AUROC | Sensitivity | Specificity | Accuracy |
|---|---|---|---|---|---|---|
| HC vs. CC | | | | | | |
| | 0.25 | 0.519 | 0.025 | 0.557 | 0.473 | 0.515 |
| | 0.50 | 0.517 | 0.024 | 0.508 | 0.516 | 0.512 |
| | 0.75 | 0.518 | 0.023 | 0.553 | 0.476 | 0.515 |
| | 1.00 | 0.517 | 0.024 | 0.571 | 0.460 | 0.515 |
| HC vs. STB | | | | | | |
| | 0.25 | 0.564 | 0.058 | 0.537 | 0.551 | 0.544 |
| | 0.50 | 0.563 | 0.063 | 0.538 | 0.549 | 0.543 |
| | 0.75 | 0.560 | 0.058 | 0.532 | 0.551 | 0.541 |

| | | | | | |
|---|---|---|---|---|---|
| 1.00 | 0.560 | 0.057 | 0.544 | 0.537 | 0.540 |
| CC vs. STB | | | | | |
| 0.25 | 0.522 | 0.045 | 0.645 | 0.387 | 0.516 |
| 0.50 | 0.521 | 0.049 | 0.634 | 0.398 | 0.516 |
| 0.75 | 0.523 | 0.047 | 0.658 | 0.385 | 0.521 |
| 1.00 | 0.517 | 0.048 | 0.651 | 0.371 | 0.511 |

Note: CC: clinical control group;  HC: healthy control group; STB: suicidal thoughts and behaviors group

### Supplemental Table S9. Classification of STB groups (parent-reported): Results of binomial penalized logistic regression analysis only including specific types of predictors

**Sociodemographic**

| Comparison | Alpha | AUROC | SD AUROC | Sensitivity | Specificity | Accuracy |
|---|---|---|---|---|---|---|
| HC vs. CC | | | | | | |
| | 0.25 | 0.555 | 0.030 | 0.542 | 0.537 | 0.539 |
| | 0.50 | 0.551 | 0.027 | 0.530 | 0.545 | 0.537 |
| | 0.75 | 0.553 | 0.029 | 0.534 | 0.540 | 0.537 |
| | 1.00 | 0.550 | 0.029 | 0.532 | 0.539 | 0.536 |
| HC vs. STB | | | | | | |
| | 0.25 | 0.612 | 0.066 | 0.573 | 0.594 | 0.584 |
| | 0.50 | 0.604 | 0.071 | 0.575 | 0.586 | 0.580 |
| | 0.75 | 0.603 | 0.069 | 0.589 | 0.579 | 0.584 |
| | 1.00 | 0.603 | 0.061 | 0.555 | 0.593 | 0.574 |
| CC vs. STB | | | | | | |
| | 0.25 | 0.550 | 0.057 | 0.571 | 0.516 | 0.543 |
| | 0.50 | 0.541 | 0.068 | 0.601 | 0.458 | 0.530 |
| | 0.75 | 0.534 | 0.067 | 0.582 | 0.477 | 0.529 |
| | 1.00 | 0.545 | 0.060 | 0.568 | 0.505 | 0.536 |

**Physical health**

| Comparison | Alpha | AUROC | SD AUROC | Sensitivity | Specificity | Accuracy |
|---|---|---|---|---|---|---|
| HC vs. CC | | | | | | |
| | 0.25 | 0.662 | 0.026 | 0.687 | 0.549 | 0.618 |
| | 0.50 | 0.663 | 0.026 | 0.689 | 0.549 | 0.619 |
| | 0.75 | 0.661 | 0.029 | 0.690 | 0.545 | 0.617 |
| | 1.00 | 0.662 | 0.025 | 0.690 | 0.546 | 0.618 |
| HC vs. STB | | | | | | |
| | 0.25 | 0.770 | 0.052 | 0.629 | 0.770 | 0.699 |
| | 0.50 | 0.767 | 0.050 | 0.625 | 0.773 | 0.699 |
| | 0.75 | 0.774 | 0.051 | 0.633 | 0.782 | 0.707 |
| | 1.00 | 0.768 | 0.058 | 0.626 | 0.777 | 0.702 |
| CC vs. STB | | | | | | |
| | 0.25 | 0.632 | 0.061 | 0.516 | 0.686 | 0.601 |
| | 0.50 | 0.635 | 0.067 | 0.517 | 0.683 | 0.600 |
| | 0.75 | 0.631 | 0.061 | 0.521 | 0.674 | 0.597 |

| Comparison | Alpha | AUROC | SD AUROC | Sensitivity | Specificity | Accuracy |
|---|---|---|---|---|---|---|
| | 1.00 | 0.644 | 0.057 | 0.517 | 0.700 | 0.608 |
| **Social environment** | | | | | | |
| Comparison | Alpha | AUROC | SD AUROC | Sensitivity | Specificity | Accuracy |
| HC vs. CC | | | | | | |
| | 0.25 | 0.602 | 0.026 | 0.668 | 0.475 | 0.571 |
| | 0.50 | 0.602 | 0.030 | 0.669 | 0.476 | 0.572 |
| | 0.75 | 0.603 | 0.031 | 0.670 | 0.478 | 0.574 |
| | 1.00 | 0.606 | 0.026 | 0.670 | 0.482 | 0.576 |
| HC vs. STB | | | | | | |
| | 0.25 | 0.710 | 0.053 | 0.551 | 0.768 | 0.659 |
| | 0.50 | 0.710 | 0.065 | 0.547 | 0.766 | 0.656 |
| | 0.75 | 0.702 | 0.063 | 0.555 | 0.753 | 0.654 |
| | 1.00 | 0.694 | 0.054 | 0.542 | 0.750 | 0.646 |
| CC vs. STB | | | | | | |
| | 0.25 | 0.602 | 0.055 | 0.477 | 0.669 | 0.573 |
| | 0.50 | 0.611 | 0.063 | 0.484 | 0.673 | 0.578 |
| | 0.75 | 0.606 | 0.063 | 0.474 | 0.683 | 0.578 |
| | 1.00 | 0.612 | 0.056 | 0.466 | 0.689 | 0.578 |
| **Clinical psychiatric** | | | | | | |
| Comparison | Alpha | AUROC | SD AUROC | Sensitivity | Specificity | Accuracy |
| CC vs. STB | | | | | | |
| | 0.25 | 0.775 | 0.056 | 0.658 | 0.751 | 0.704 |
| | 0.50 | 0.777 | 0.053 | 0.655 | 0.761 | 0.708 |
| | 0.75 | 0.769 | 0.052 | 0.661 | 0.748 | 0.705 |
| | 1.00 | 0.782 | 0.049 | 0.661 | 0.766 | 0.713 |
| **Cognition** | | | | | | |
| Comparison | Alpha | AUROC | SD AUROC | Sensitivity | Specificity | Accuracy |
| HC vs. CC | | | | | | |
| | 0.25 | 0.594 | 0.028 | 0.588 | 0.555 | 0.572 |
| | 0.50 | 0.594 | 0.028 | 0.586 | 0.559 | 0.572 |
| | 0.75 | 0.592 | 0.024 | 0.586 | 0.553 | 0.570 |
| | 1.00 | 0.591 | 0.028 | 0.586 | 0.552 | 0.569 |
| HC vs. STB | | | | | | |
| | 0.25 | 0.647 | 0.055 | 0.587 | 0.628 | 0.608 |
| | 0.50 | 0.640 | 0.063 | 0.582 | 0.623 | 0.603 |

| | | | | | | |
|---|---|---|---|---|---|---|
| | 0.75 | 0.641 | 0.070 | 0.584 | 0.605 | 0.594 |
| | 1.00 | 0.642 | 0.066 | 0.584 | 0.624 | 0.604 |
| CC vs. STB | | | | | | |
| | 0.25 | 0.551 | 0.070 | 0.584 | 0.485 | 0.534 |
| | 0.50 | 0.559 | 0.077 | 0.558 | 0.530 | 0.544 |
| | 0.75 | 0.555 | 0.061 | 0.573 | 0.509 | 0.541 |
| | 1.00 | 0.550 | 0.061 | 0.573 | 0.496 | 0.534 |

**Neuroimaging**

| Comparison | Alpha | AUROC | SD AUROC | Sensitivity | Specificity | Accuracy |
|---|---|---|---|---|---|---|
| HC vs. CC | | | | | | |
| | 0.25 | 0.508 | 0.020 | 0.574 | 0.436 | 0.505 |
| | 0.50 | 0.513 | 0.024 | 0.576 | 0.439 | 0.507 |
| | 0.75 | 0.506 | 0.019 | 0.625 | 0.379 | 0.502 |
| | 1.00 | 0.510 | 0.026 | 0.546 | 0.469 | 0.507 |
| HC vs. STB | | | | | | |
| | 0.25 | 0.522 | 0.060 | 0.557 | 0.470 | 0.513 |
| | 0.50 | 0.530 | 0.068 | 0.599 | 0.445 | 0.522 |
| | 0.75 | 0.527 | 0.055 | 0.551 | 0.491 | 0.521 |
| | 1.00 | 0.514 | 0.050 | 0.548 | 0.468 | 0.508 |
| CC vs. STB | | | | | | |
| | 0.25 | 0.507 | 0.046 | 0.579 | 0.425 | 0.502 |
| | 0.50 | 0.509 | 0.051 | 0.606 | 0.410 | 0.508 |
| | 0.75 | 0.502 | 0.052 | 0.620 | 0.372 | 0.496 |
| | 1.00 | 0.504 | 0.042 | 0.643 | 0.364 | 0.504 |

**Genetic**

| Comparison | Alpha | AUROC | SD AUROC | Sensitivity | Specificity | Accuracy |
|---|---|---|---|---|---|---|
| HC vs. CC | | | | | | |
| | 0.25 | 0.529 | 0.026 | 0.560 | 0.486 | 0.523 |
| | 0.50 | 0.525 | 0.025 | 0.561 | 0.480 | 0.521 |
| | 0.75 | 0.524 | 0.025 | 0.540 | 0.499 | 0.519 |
| | 1.00 | 0.527 | 0.024 | 0.543 | 0.503 | 0.523 |
| HC vs. STB | | | | | | |
| | 0.25 | 0.549 | 0.058 | 0.574 | 0.489 | 0.532 |
| | 0.50 | 0.545 | 0.070 | 0.568 | 0.495 | 0.531 |
| | 0.75 | 0.553 | 0.058 | 0.543 | 0.527 | 0.535 |

| | | | | | |
|---|---|---|---|---|---|
| 1.00 | 0.541 | 0.058 | 0.526 | 0.527 | 0.526 |
| CC vs. STB | | | | | |
| 0.25 | 0.537 | 0.068 | 0.575 | 0.469 | 0.522 |
| 0.50 | 0.533 | 0.062 | 0.570 | 0.472 | 0.521 |
| 0.75 | 0.533 | 0.058 | 0.550 | 0.496 | 0.523 |
| 1.00 | 0.529 | 0.056 | 0.515 | 0.521 | 0.518 |

Note: CC: clinical control group; HC: healthy control group; STB: suicidal thoughts and behaviors group
